# Immune responses in COVID-19 respiratory tract and blood reveal mechanisms of disease severity

**DOI:** 10.1101/2021.09.01.21262715

**Authors:** Wuji Zhang, Brendon Y. Chua, Kevin J. Selva, Lukasz Kedzierski, Thomas M. Ashhurst, Ebene R. Haycroft, Suzanne K. Shoffner, Luca Hensen, David F. Boyd, Fiona James, Effie Mouhtouris, Jason C. Kwong, Kyra Y. L. Chua, George Drewett, Ana Copaescu, Julie E. Dobson, Louise C. Rowntree, Jennifer R. Habel, Lilith F. Allen, Hui-Fern Koay, Jessica A. Neil, Matthew Gartner, Christina Y. Lee, Patiyan Andersson, Torsten Seemann, Norelle L. Sherry, Fatima Amanat, Florian Krammer, Sarah L. Londrigan, Linda M. Wakim, Nicholas J.C. King, Dale I. Godfrey, Laura K. Mackay, Paul G. Thomas, Suellen Nicholson, Kelly B. Arnold, Amy W. Chung, Natasha E. Holmes, Olivia C. Smibert, Jason A. Trubiano, Claire L. Gordon, Thi H.O. Nguyen, Katherine Kedzierska

**Author notes:** Corresponding authors: Katherine Kedzierska, Oanh Nguyen; Claire Gordon, Jason Trubiano. Authors contributed equally to this study (Jason A. Trubiano, Claire L. Gordon, Thi H.O. Nguyen, Katherine Kedzierska).

## Abstract

Although the respiratory tract is the primary site of SARS-CoV-2 infection and the ensuing immunopathology, respiratory immune responses are understudied and urgently needed to understand mechanisms underlying COVID-19 disease pathogenesis. We collected paired longitudinal blood and respiratory tract samples (endotracheal aspirate, sputum or pleural fluid) from hospitalized COVID-19 patients and non-COVID-19 controls. Cellular, humoral and cytokine responses were analysed and correlated with clinical data. SARS-CoV-2-specific IgM, IgG and IgA antibodies were detected using ELISA and multiplex assay in both the respiratory tract and blood of COVID-19 patients, although a higher receptor binding domain (RBD)-specific IgM and IgG seroconversion level was found in respiratory specimens. SARS-CoV-2 neutralization activity in respiratory samples was detected only when high levels of RBD-specific antibodies were present. Strikingly, cytokine/chemokine levels and profiles greatly differed between respiratory samples and plasma, indicating that inflammation needs to be assessed in respiratory specimens for the accurate assessment of SARS-CoV-2 immunopathology. Diverse immune cell subsets were detected in respiratory samples, albeit dominated by neutrophils. Importantly, we also showed that dexamethasone and/or remdesivir treatment did not affect humoral responses in blood of COVID-19 patients. Overall, our study unveils stark differences in innate and adaptive immune responses between respiratory samples and blood and provides important insights into effect of drug therapy on immune responses in COVID-19 patients.

## INTRODUCTION

Symptoms of SARS-CoV-2 infection, known as coronavirus disease 2019 (COVID-19), vary from asymptomatic or mild disease to critical illness, including respiratory failure and death^1^. Global efforts focused on developing new drugs and vaccines. While vaccines showed immunogenicity and safety towards SARS-CoV-2^2, 3, 4^, effects of drug treatments remain controversial. Dexamethasone, a synthetic glucocorticoid drug, can lower the 28-day mortality rate in COVID-19 patients receiving oxygen support, prolong ventilator-free days and improve oxygen partial pressure to fractional inspired oxygen (PaO_2_/FiO_2_) ratio compared to placebo or standard care^5, 6, 7^. However, SARS-CoV-2 RNA can be detected for longer in patients receiving glucocorticoid treatment^8^. Treatment with remdesivir, a nucleoside analogue inhibiting RNA-dependent RNA polymerase (RdRp) in COVID-19 can shorten time to recovery and provide better clinical outcomes^9, 10, 11^. However, the effects of dexamethasone and/or remdesivir on humoral and cellular immune responses are unclear.

Immunity towards SARS-CoV-2 infection has been studied, predominantly in peripheral blood. While robust, broad and transient immune responses precede patients’ recovery in non-severe cases^12, 13, 14, 15^, severe COVID-19 can be associated with exuberant cytokine responses, hyperactivation of innate immune cells, reduced T-cell numbers^12, 14, 16, 17^ and high titres of SARS-CoV-2-specific antibodies^16^. In contrast, immune responses in the respiratory tract are understudied. High levels of IL-6, IL-10, monocyte chemoattractant protein (MCP)-1, macrophage inflammatory protein (MIP)-1α and MIP-1β are detected in bronchoalveolar lavage fluid (BALF) of COVID-19 patients, indicating inflammatory environment with high monocyte chemoattractants^18, 19^. While IFN-α and IFN-β were undetectable, IL-10, IL-17A, IL-18 were variably detected in COVID-19 BALF, with higher RNA and/or protein levels of IL-6, MCP-1, IL-33 and lower IL-6 receptor (IL-6R) observed in COVID-19 BALF compared to healthy BALF^19, 20^. Granulocytes and monocytes/macrophages dominate in COVID-19 airways, especially intermediate (CD14^+^CD16^+^) and non-classical (CD14^-^CD16^+^) monocytes^18, 21^. Conversely, low frequencies of T-cells were detected in COVID-19 airways with increased expression of activation markers CD38/HLA-DR and a tissue-resident phenotype^18, 19^. Increased frequency of activated T-cells in the airway is associated with improved survival^18^.

To dissect the breadth of immune responses during SARS-CoV-2 infection in the respiratory tract compared to those detected in blood, we collected paired longitudinal blood and respiratory samples from hospitalised COVID-19 patients and non-COVID-19 controls to investigate innate, adaptive and humoral immunity. While discordant cytokine levels were detected in respiratory samples across COVID-19 patients, higher IgM and IgG seroconversion was found in respiratory samples compared to paired blood. While higher frequencies of neutrophils and effector memory (EM)-like CD4^+^ and CD8^+^ T-cells were found in COVID-19 respiratory samples, higher antibody levels also correlated with activated cellular immunity. Elevated soluble IL-6R alpha (sIL-6Rα) levels and more robust humoral responses were detected in blood of patients with severe disease, and importantly, dexamethasone (with/without) remdesivir therapy did not reduce immune responses in COVID-19 patients. Overall, our study unveils stark differences in innate and adaptive immune responses between respiratory and blood samples of COVID-19 patients and provides insights into potential biomarkers and immunotherapies for severe COVID-19.

## RESULTS

### DRASTIC patient cohort

We recruited 66 patients hospitalized at Austin Hospital (Victoria, Australia) with suspected COVID-19 into the pre<u>D</u>icto<u>R</u>s of dise<u>A</u>se <u>S</u>everity in cri<u>T</u>ically Ill <u>C</u>OVID-19 (DRASTIC) cohort prior to their PCR result (Fig. 1a, Supplementary Table 1). Sixty patients had PCR-confirmed SARS-CoV-2 infection, including 43 ward patients (34.9% requiring non-invasive oxygen support) and 17 patients requiring admission to the ICU (35.3% requiring mechanical ventilation; 58.8% requiring non-invasive oxygen support), while 6 patients were SARS-CoV-2-negative and respiratory IgG RBD ELISA-negative (1 ward; 5 ICU patients; Supplementary Table 2). 12 ward patients and six ICU patients were on dexamethasone treatment, while 8 ward patients and 10 ICU patients were on dexamethasone (with/without remdesivir) treatment. The median age of COVID-19 patients was 58 years (range 22-90) and 46.7% were females. The median age was 58 years within both ward and ICU patients (Supplementary Table 2).

**Fig. 1.**
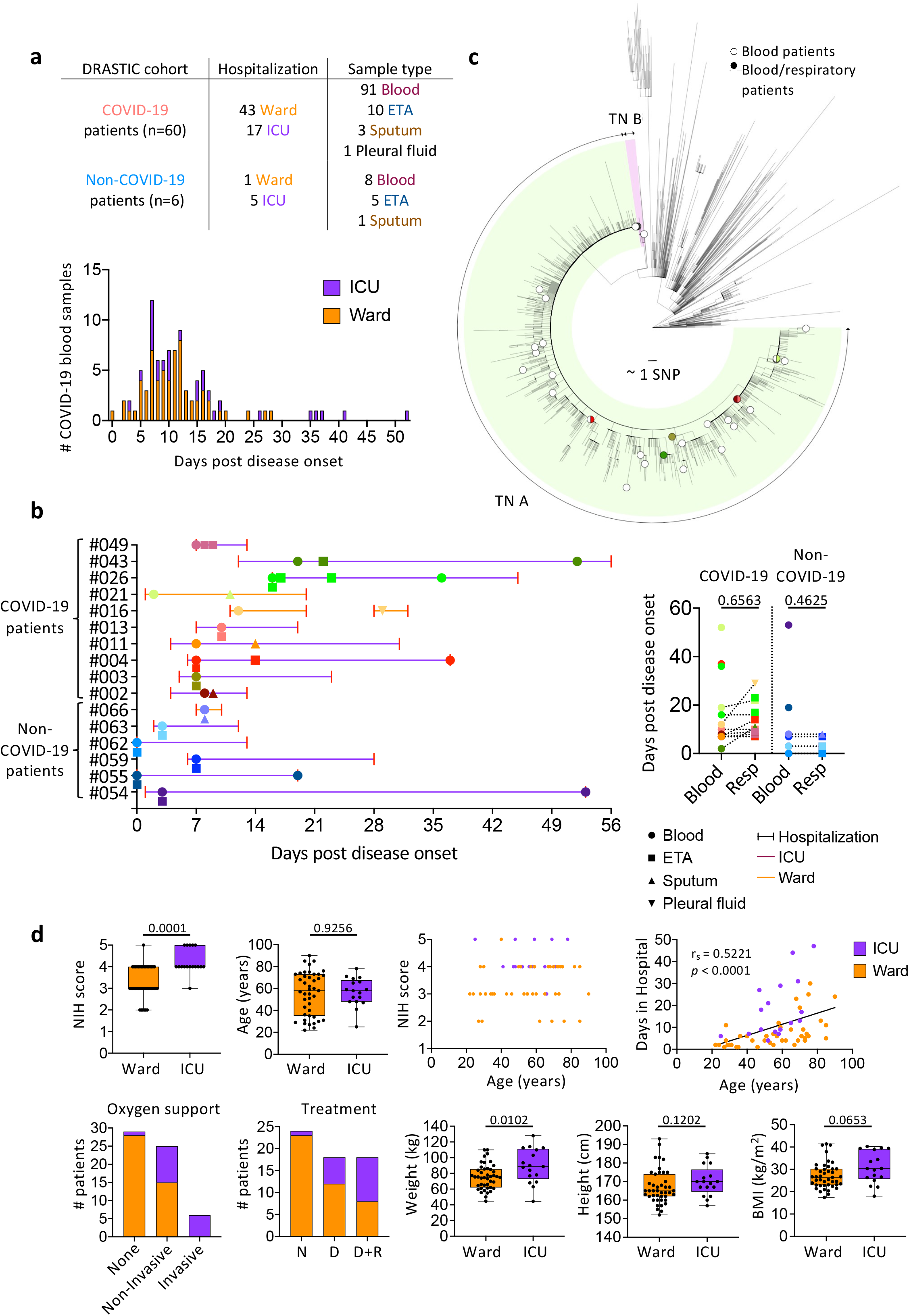
Pre<u>D</u>icto<u>R</u>s of dise<u>A</u>se <u>S</u>everity in cri<u>T</u>ically <u>I</u>ll <u>C</u>OVID-19 patient (DRASTIC) cohort. **a** Number of patients recruited in the DRASTIC cohort and samples collected (top panel) and days post disease onset of COVID-19 blood samples (bottom panel). **b** Collection timepoints of respiratory samples (endotracheal tube aspirate (ETA), sputum, and pleural fluid) and paired blood samples from the same patients (left panel). Comparison of days post disease onset between blood and respiratory samples for COVID-19 and non-COVID-19 patients (right panel) using Mann-Whitney test. **c** Maximum likelihood phylogenetic tree of SARS-CoV-2 sequences from Victoria from 28 January 2020 to 28 October 2020 (including context sequences from rest of Australia and New Zealand). Phylogenetic tree includes randomly subsampled sequences from transmission networks (TN) A and TN B in Victoria, with a total number of 10941 and 145 cases respectively. The outermost tip of each radial line represents a single sequence; the sum of each radial line between two tips represents the genetic distance between two sequences. Each radial stepwise progression represents approximately one single nucleotide polymorphism (SNP). Sequences from study patients are shown as open circles (patient with blood samples only) or solid-coloured circles (patients with blood and respiratory samples, same colours as in section b). Half-filled circles are used when samples are located close to each other. **d** Distribution of clinical data in ward and intensive care unit (ICU) COVID-19 patients. Box and bars indicate first and third quartiles, and range respectively. Statistical significance was determined with the Fisher’s exact test for the National Institutes of Health (NIH) score, and Mann-Whitney test for age, weight, height, and body weight index (BMI). Correlation was determined with Spearman’s correlation.

97% of blood samples were collected at hospital admission (Visit 1/V1) or discharge (V7; n=62 and n=35 out of 99 blood samples, respectively), with others collected during hospitalization (V2:ICU admission, V3:acute respiratory distress syndrome/cytokine release syndrome diagnosis, V5:24-48 hours post drug therapy, V6:7-14 days post drug therapy). While 83 out of 91 blood samples from 53 COVID-19 patients were collected within 24 days post disease onset (median 10 days, range 0-24 days) (Fig. 1a), 6 patients had longer hospital stays (median 32 days, range 26-52 days) (Supplementary Table 1).

Additionally, 20 respiratory samples (15 endotracheal aspirate (ETA), 4 sputum, 1 pleural fluid) were obtained from 16 DRASTIC patients (10 COVID-19 patients and 6 non-COVID-19 patients) at 0 days (median) after first blood collection (range 0-9 days), with no significant differences between time of blood versus respiratory specimen collection (Fig. 1b). The pleural fluid was collected at re-admission (Fig. 1b). Respiratory samples were obtained from 2 ward COVID-19 patients and 8 ICU COVID-19 patients. While ETA samples were collected from intubated patients, sputum samples were collected from non-intubated patients. Respiratory samples were also collected from 5 non-COVID-19 hospitalized ICU patients requiring invasive ventilation and 1 ward patient requiring non-invasive oxygen support (Fig. 1a, b).

SARS-CoV-2 genome sequence data, available from 41 out of 60 COVID-19 patients, showed that patients were infected with SARS-CoV-2 viruses belonging to the transmission network (TN)-A, representing a highly clonal and dominant network during the second wave of COVID-19 epidemic in Victoria^22^, except for 1 patient belonging to TN-B (DRASTIC-009) (Fig. 1c).

### ICU admission associated with higher NIH severity score, oxygen therapy, drug treatment and weight

Disease severity within the DRASTIC cohort was stratified according to whether COVID-19 patients were hospitalized in the ward or ICU. COVID-19 patients were also graded according to the NIH severity score of 1-5 according to their symptoms (Supplementary Table 3). ICU patients had significantly higher NIH scores compared to ward patients (*p*=0.0001, Fig. 1d) and more ICU patients received oxygen support (*p*<0.0001) and drug treatments, either dexamethasone alone or dexamethasone with remdesivir (*p*=0.0009) (Supplementary Table 2). Interestingly, ICU patients also had significantly increased body weight (*p*=0.0102). Age correlated with the length of hospital stay (*p*<0.0001, Fig. 1d), but no differences in age, gender, height, ethnicity, immunosuppressant drugs or smoking were observed between ICU and ward patients (Fig. 1d, Supplementary Table 3).

### Inflammatory cytokines in respiratory samples are not reflective of plasma inflammation

There are scarce data on inflammatory milieu in COVID-19 respiratory specimens. To determine cytokine/chemokine levels and composition in respiratory samples compared to paired plasma, we measured cytokines/chemokines (IL-1β, IFN-⍰2, IFN-γ, TNF, MCP-1, IL-6, IL-8, IL-10, IL-12p70, IL-17A, IL-18, IL-23, IL-33), sIL-6R⍰ and an extracellular matrix protein “disintegrin and metalloproteinase with thrombospondin motifs-4” (ADAMTS4)^23^. Cytokine levels varied across both COVID-19 patients in respiratory samples and between paired respiratory and plasma samples. Five COVID-19 ICU patients (DRASTIC-002, -003, -004, -011, -013) had no detectable cytokines across 13 cytokines/chemokines, while high IL-18 levels were detected in plasma of DRASTIC-002, -003, -004 and -013 patients (Fig. 2a). Amongst the remaining COVID-19 patients, greatly elevated levels of inflammatory cytokines/chemokines were detected in respiratory samples, with concentrations being 60× (MCP-1), 400× (IL-6) and 780× (IL-8) higher than in plasma for DRASTIC-021, -026, -043, -049. While IL-18 dominated in plasma, IL-6, IL-8 and MCP-1 were most prevalent in respiratory samples in patients with high cytokines/chemokines (Fig. 2a, Supplementary Fig. 1a).

**Fig. 2.**
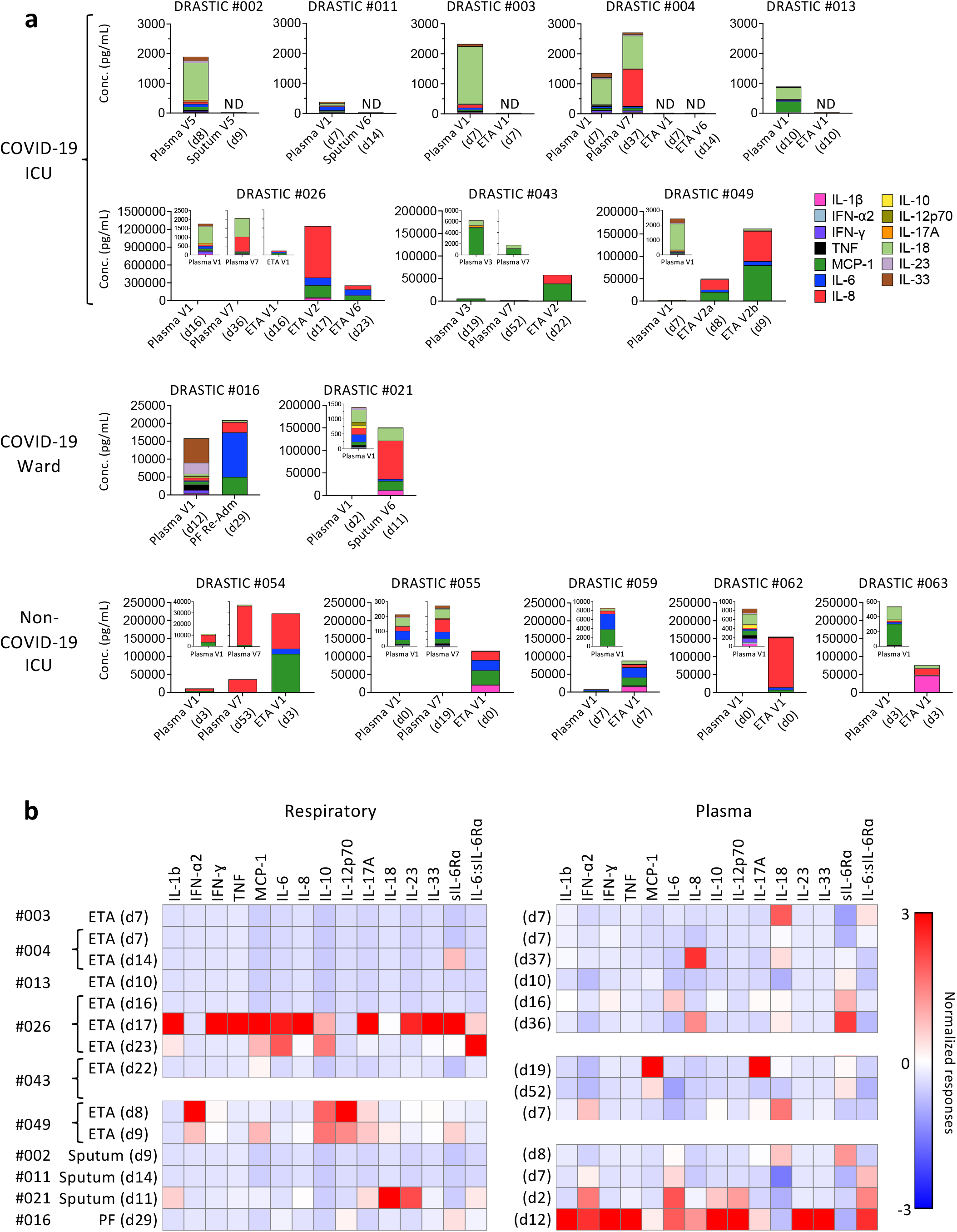
Discordant levels of cytokines and chemokines in COVID-19 respiratory samples compared to paired plasma samples. **a** Absolute concentrations of 13 cytokines and chemokines (IL-1β, IFN-⍰2, IFN-γ, TNF, MCP-1, IL-6, IL-8, IL-10, IL-12p70, IL-17A, IL-18, IL-23, and IL-33) in each individual with respiratory and plasma samples. **b** Standardized levels of cytokines/chemokines for COVID-19 respiratory and plasma samples separately. Red color indicates higher cytokine/chemokine levels. PF, pleural fluid; Re-Adm, re-admission.

This disparity was also reflected when cytokine and sIL-6R⍰ levels were standardized separately for respiratory and plasma samples. With the red color indicating higher cytokine levels, donors with high cytokine concentration in the respiratory samples did not necessarily display high levels of the same cytokine in their plasma (Fig. 2b). For instance, patient DRASTIC-049 displayed higher IFN-⍰2, IL-10, IL-12p70 and IL-17A in ETA (d8) than other COVID-19 respiratory samples, while the plasma (d7) level of IFN-⍰2 was also higher, DRASTIC-049 had higher plasma level of IL-18 but not IL-10, IL-12p70 or IL-17A (Fig 2b).

In contrast, respiratory samples from non-COVID-19 ICU patients showed significantly higher levels of several cytokines, including IL-6, IL-6:sIL-6Rα ratio, IL-1β, IL-8, IL-10, IL-18, IL-23, TNF, MCP-1 compared to matched plasma samples (7-440× higher) (Fig. 2a, Supplementary Fig. 1b), reflecting consistent levels of those cytokines across non-COVID-19 ICU patients. sIL-6R⍰ in non-COVID-19 respiratory samples was lower compared to plasma, while comparable to COVID-19 respiratory samples (Supplementary Fig. 1b).

Overall, while the inflammatory cytokine/chemokine levels were excessively higher in respiratory fluid compared to plasma in some COVID-19 patients, they were highly variable across COVID-19 patients, indicating that the plasma inflammatory milieu does not reflect the airway inflammation and that hospitalized/ICU COVID-19 patients should be monitored for inflammation in airways to understand disease severity and potential benefits of immunomodulatory treatments.

### High RBD-specific IgM and IgG seroconversion in COVID-19 respiratory samples

SARS-CoV-2-specific antibodies in respiratory samples are relatively unexplored. We measured SARS-CoV-2 RBD-specific IgM, IgG and IgA antibodies in paired respiratory and blood samples using RBD-ELISA and surrogate virus neutralisation test (sVNT) (Fig. 3). Compared to non-COVID-19, COVID-19 patients displayed higher levels of RBD IgM (*p*=0.0019) and IgG (*p*=0.0004) in respiratory samples (Fig. 3a, b), but not RBD IgA levels, which was possibly due to either technical issues or cross-reactivity of IgA antibodies (Fig. 3bi). Moreover, comparable titres of RBD IgM and IgG were found in COVID-19 respiratory samples and plasma (Fig. 3bii). In 3 DRASTIC patients (−004, -026, -049) with sequential ETAs collected, RBD-specific IgM, IgG, and IgA antibody titres increased in both plasma and ETA samples over time (Fig. 3c).

**Fig. 3.**
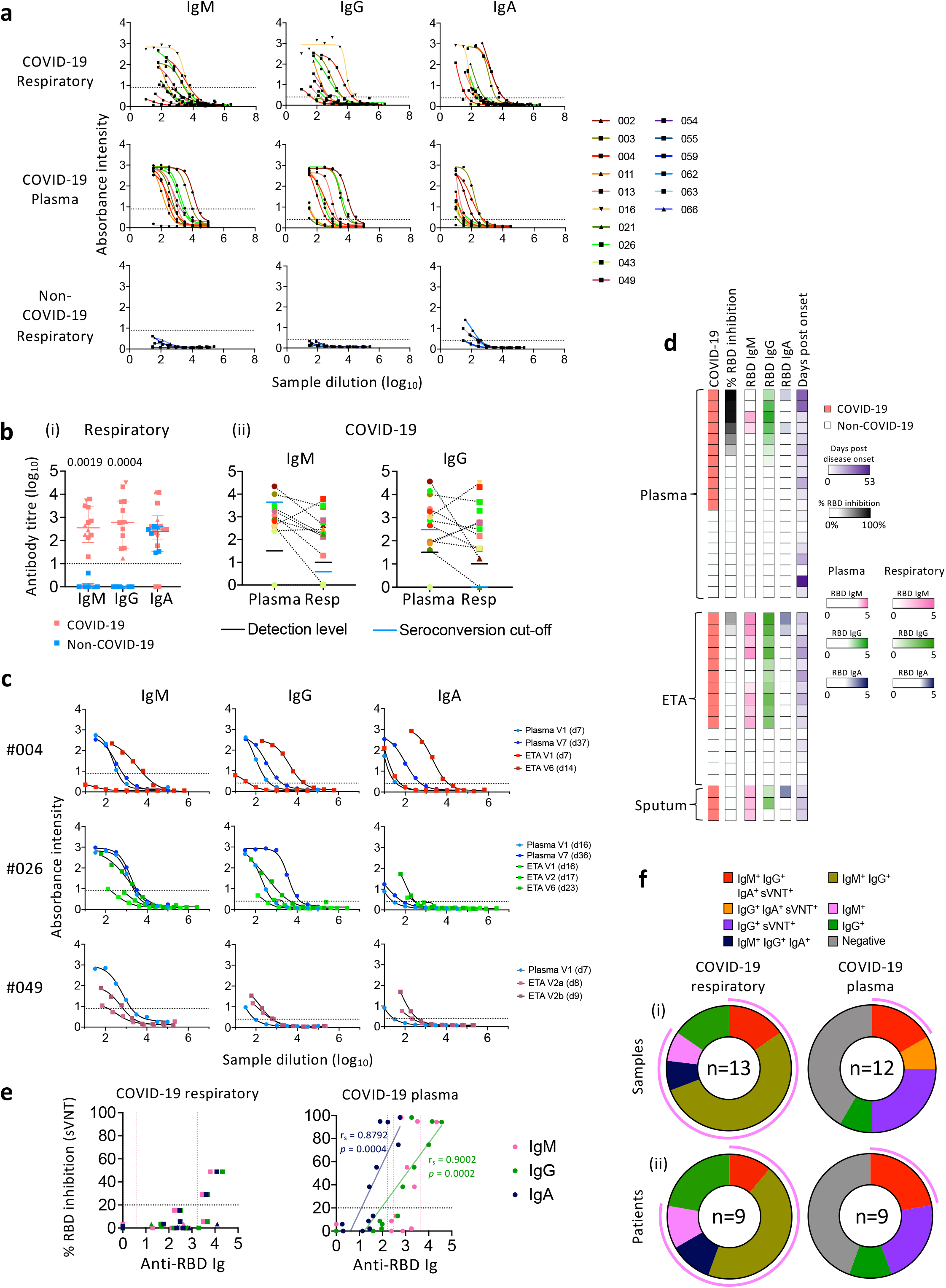
Higher anti-RBD IgM seroconversion rate in respiratory samples compared to paired blood samples of COVID-19 patients. **a** ELISA titration curves against the SARS-CoV-2 receptor-binding domain (RBD) for IgM, IgG, and IgA in COVID-19 respiratory and paired blood samples and non-COVID-19 respiratory samples. Dotted lines within each graph indicates the cut-off used to determine end-point titres. **b** End-point titres of SARS-CoV-2 RBD antibodies between (i) respiratory samples of COVID-19 and non-COVID-19 patients, and (ii) plasma and respiratory samples of COVID-19 patients. (i) Bars indicate median with interquartile range. Dotted line indicates the detection level. (ii) Dotted lines connect the most closely matched plasma and respiratory samples from each patient. Statistical significance was determined with Mann-Whitney test. **c** ELISA titration curves against the SARS-CoV-2 RBD for 3 COVID-19 patients with serial respiratory samples. **d** Heatmap of percentage (%) inhibition tested by surrogate virus neutralization test (sVNT) and anti-RBD ELISA titres. **e** Correlation between anti-RBD antibody titres and (%) sVNT inhibition. Correlation was determined with Spearman’s correlation. **f** Number of (i) samples and (ii) patients with seroconverted anti-RBD IgM, IgG, IgA and positive % sVNT inhibition. Pink curved lines surrounding the donut graphs indicate the samples/patients with seroconverted IgM. Earliest samples were used for each patient when determining seroconversion which was defined as average titre +2×SD of non-COVID-19 samples. Positive % sVNT inhibition was defined as % sVNT inhibition ≥ 20%.

Using sVNT, two ETA samples from DRASTIC-003 and -004 (with low cytokine levels) had detectable neutralizing activity, associated with high levels of RBD-specific IgM, IgG, and IgA antibodies (Fig. 3d, e). Neutralizing activity was not detected in the remaining respiratory samples at the acute time-points. Plasma samples with high neutralizing activity had high levels of all three IgM, IgG and IgA isotypes of RBD-specific antibodies, and anti-RBD IgG and IgA positively correlated with the neutralizing activity (*p*=0.0002; *p*=0.0004 respectively, Fig. 3d, e). Seroconverted levels of RBD-specific IgM and IgG antibodies were detected in the majority of COVID-19 respiratory samples (10/13) and patients (6/9) at 77% and 67% (Fig. 3f), suggesting prominence of RBD-specific IgM and IgG in respiratory samples during acute COVID-19.

### High prevalence of SARS-CoV-2-NP-specific IgM antibodies in respiratory samples

While anti-RBD antibodies are essential for the neutralization of SARS-CoV-2^24^, non-neutralizing antibodies are important role in antiviral immunity^25^. To understand in-depth antibody profiles and cross-reactivity in respiratory samples, we adapted a multiplex bead array assay^25^ (Supplementary Table 5). Antibodies targeting RBD, S proteins, NP of SARS-CoV-2, SARS-CoV-1 and human coronaviruses (229E, NL63, OC43, HKU1) along with isotypes/subclasses (IgM, IgG, IgG1-4, IgA1-2), binding with FcγR (FcγR2aH, FcγR2aR, FcγR2b, FcγR3aV, FcγR3aF) and C1q, totaling 315 features, were assessed in 14 COVID-19 and 5 non-COVID-19 respiratory samples, and paired plasma. Intermediate to high antibody levels across different SARS-CoV-2 antigens and isotypes were detected in a subset of COVID-19 respiratory and plasma samples (Fig. 4a), especially in patients who lacked inflammatory cytokines in their respiratory samples (DRASTIC-002, -003, -004, -011, -013). Conversely, patients with low anti-SARS-CoV-2 antibodies (DRASTIC-021, -043, -049) could still have variable antibody responses towards other human coronavirus in plasma and/or respiratory samples (Supplementary Fig. 2a).

**Fig. 4.**
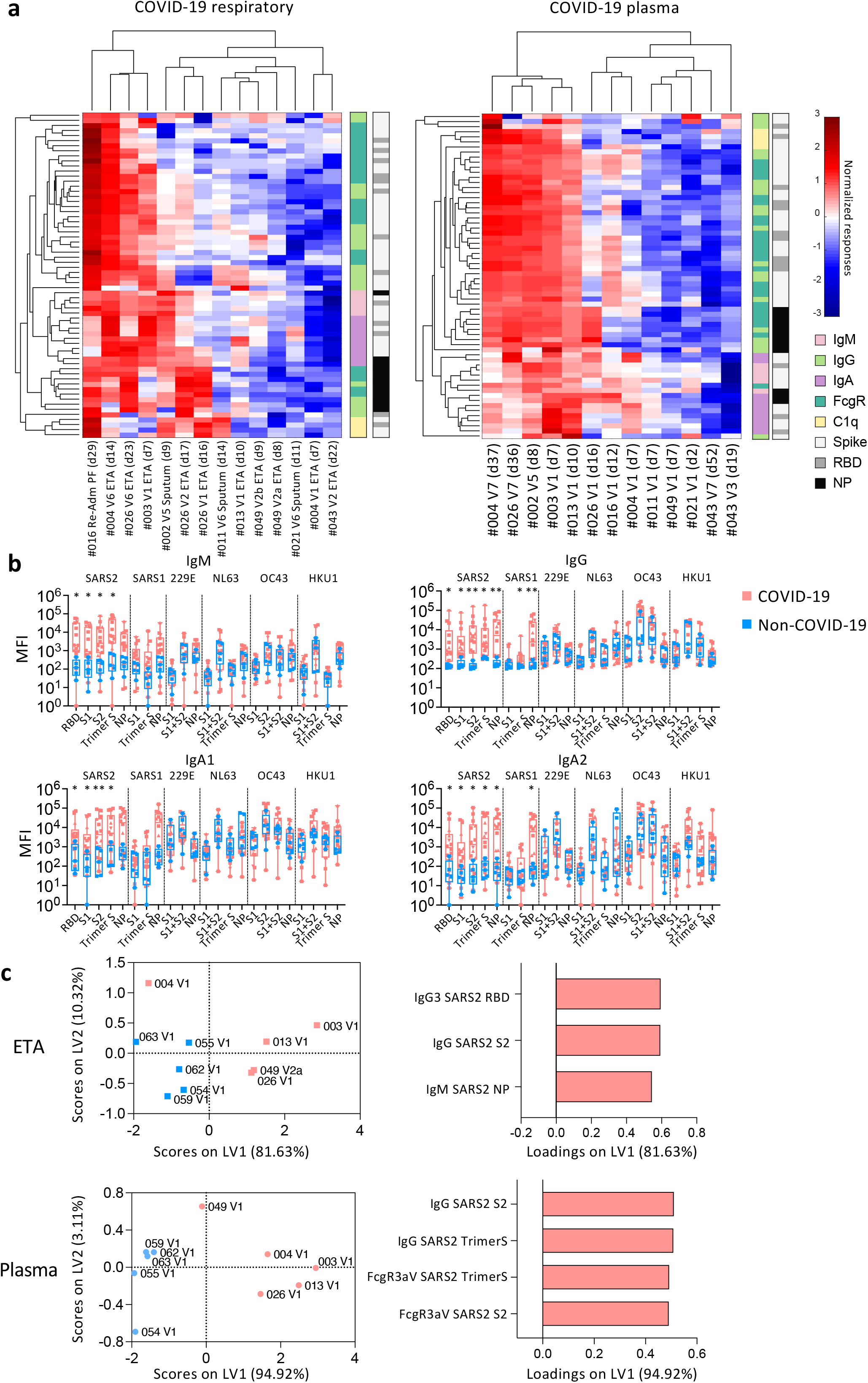
Higher anti-SARS2-NP IgM in COVID-19 ETA than non-COVID-19 ETA. **a** Heatmaps with unsupervised clustering of SARS-CoV-2-specific antibodies in COVID-19 respiratory and plasma samples. **b** median fluorescence intensity of IgM, IgG, IgA1, and IgA2 antibodies between COVID-19 and non-COVID-19 respiratory samples. Statistical significance was determined with Mann-Whitney test. **c** Partial Least-Squares Discriminant Analysis (PLSDA) scores and loading plots of ETA and plasma from five COVID-19 and five non-COVID-19 patients with the smallest difference in days post disease onset between ETA and plasma samples.

When comparing COVID-19 and non-COVID-19 respiratory samples, high levels of SARS-CoV-2-specific IgM, IgG, IgA1 and IgA2 were detected in COVID-19 (Fig. 4b). While low level SARS-CoV-1 IgG and IgA2 levels were detected, no significant differences in antibodies against other human coronaviruses (229E, NL63, OC43, HKU1) were found (Fig. 4b). IgG1 and IgG3 were the most prominent subclasses (Supplementary Fig. 2b). SARS-CoV-2 antibodies with FcγR binding abilities were detected at low levels in COVID-19 respiratory samples (Supplementary Fig. 2b).

To investigate the most prominent antibody features different between COVID-19 and non-COVID-19 patients, Partial Least-Squares Discriminant Analysis (PLSDA) was performed (Fig. 4c). As few as 3 antibody features were sufficient to separate COVID-19 and non-COVID-19 ETA, with higher SARS-CoV-2-specific IgG and IgM in COVID-19 ETA, consistent with higher anti-RBD IgG and IgM in ELISA (Fig. 3b, 4c). In contrast, higher SARS-CoV-2-S-specific IgG antibodies and antibodies with FcγR binding activities were strongly featured in COVID-19 plasma compared to non-COVID-19 plasma (Fig. 4c).

Overall, high anti-SARS-CoV-2 antibodies were detected in COVID-19 airways. While anti-SARS-CoV-2 IgM and IgG were prominent in COVID-19 airways, anti-SARS-CoV-2 antibodies with FcγR binding activities were more prevalent in COVID-19 plasma.

### Increasing cellular infiltrates in respiratory specimens during disease progression

To determine cellular immunity in respiratory specimens of COVID-19 patients, samples underwent multi-parameter flow cytometry and analysis using the Spectre R package^26^. Cells were clustered using Flow Self-Organizing Map (FlowSOM)^27^ and plotted using Fast Interpolation-based t-distributed Stochastic Neighbour Embedding (FIt-SNE)^28^. Two flow cytometry panels were used to ensure accurate profiling of myeloid and lymphoid cell populations (Supplementary Fig. 3a, 4a, 4b, Supplementary Table 6).

Clustering of respiratory samples in the myeloid panel revealed that CD66b^+^neutrophils dominated, with varying levels of CD16 expression (Fig. 5a). CD14^+^macrophages and CD4^+^ and CD8^+^ T-cells were also detected but at lower frequencies. While the cellular component was variable across samples, CD16^hi^ and CD16^lo^ neutrophils were present in all COVID-19 patients apart from DRASTIC-043 (BMT recipient; Fig. 5a, Supplementary Fig. 4c). Although there were only two COVID-19 patients (DRASTIC-026, - 049) with multiple ETA samples, we still observed an increase in cellular infiltrates was detected over time, including CD16^lo^ neutrophils (Fig. 5b). In the respiratory specimens of 6 non-COVID-19 patients, lower levels of neutrophils and macrophages were detected (Supplementary Fig. 4c). DRASTIC-059 had a large population of CD16^-^ neutrophils, while a high frequency of CD16^lo^neutrophils was detected in blood, indicating a dominant immature neutrophil population in this patient (Supplementary Fig. 4c, e).

**Fig. 5.**
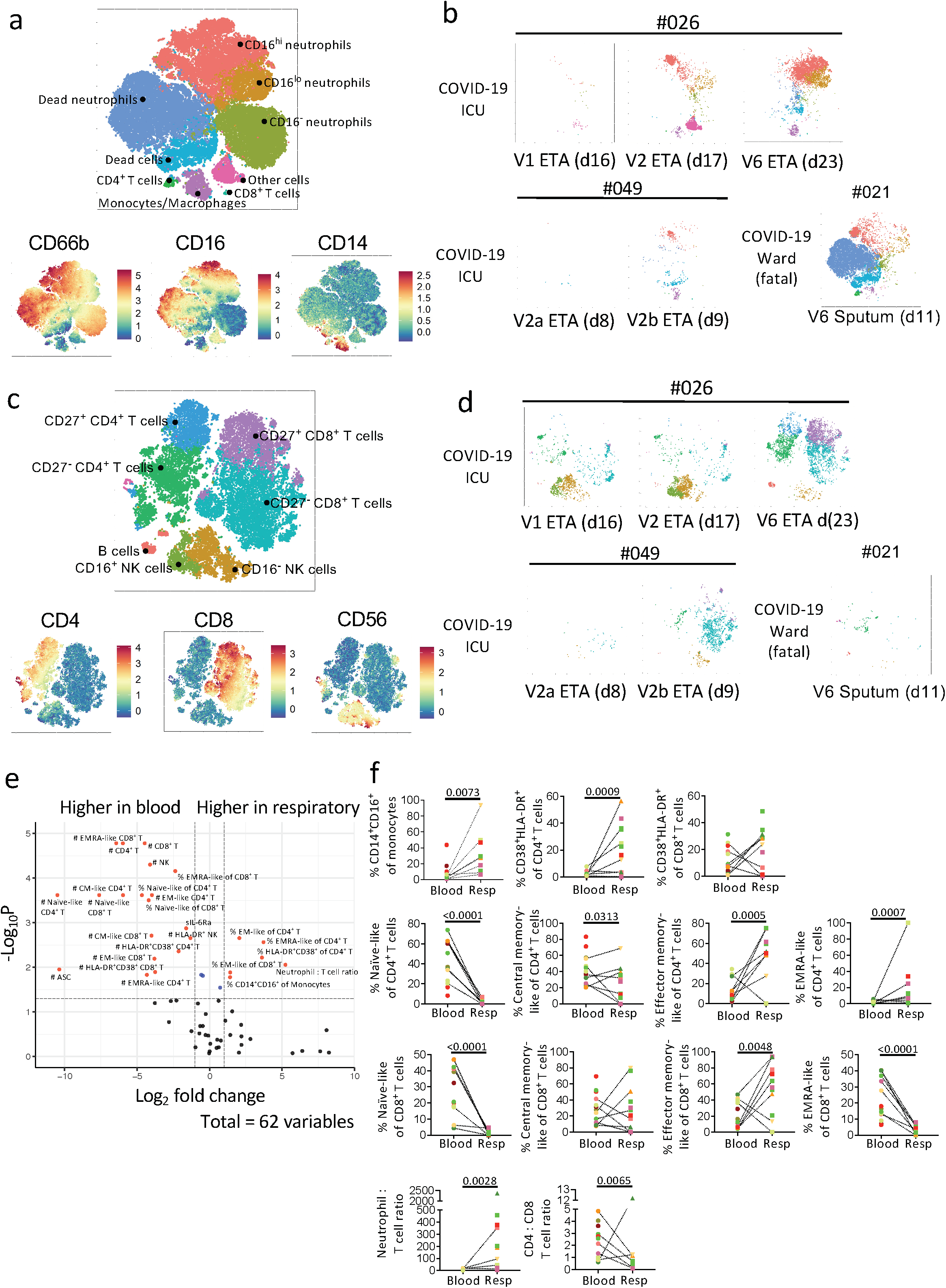
Higher frequencies of activated immune cells and EM-like CD4^+^ and CD8^+^ T cells in COVID-19 respiratory compared to paired blood samples. Flow Self-Organizing Map (FlowSOM) analyses of cellular content in the respiratory tract. **a**-**d** Metacluster of cells and expression level of the **a** myeloid antibody panel and **c** lymphocyte antibody panel. Multiple respiratory samples from patients 026 and 049 as well as one fatal patient 021 were shown as example **b, d. e** Volcano plot showing fold difference of 62 immunological features between paired respiratory and blood samples. **f** Comparisons of cellular immune features between respiratory and paired blood samples. Statistical significance was determined with Mann-Whitney test. Dotted lines connect the most closely matched plasma and respiratory samples from each patient.

After excluding neutrophils and monocytes/macrophages in respiratory samples, CD8^+^ T-cells were the major population of lymphocytes, with varying levels of CD4^+^ T-cells and natural killer (NK) cells (Fig. 5c, Supplementary Fig. 4d). Increasing infiltrates of T-cells over time were found in patients 026 and 049 (Fig. 5d), similar to neutrophils. Interestingly, in patient 026, the lymphocyte population was dominated by NK cells early (V1 and V2) and T-cells gradually infiltrated and dominated over time (V6). Low lymphocyte levels were detected in fatal patient-021.

A volcano plot was generated to determine fold-differences in immunological features between respiratory and plasma samples (Fig. 5e, f). While cell numbers were higher in blood, higher frequencies of intermediate (CD14^+^CD16^+^) monocytes/macrophages, activated (HLADR^+^CD38^+^) and EM-like (CD27^-^CD45RA^-^) CD4^+^ and CD8^+^ T-cells were found in COVID-19 respiratory specimens compared to blood. Respiratory specimens had higher neutrophil to T-cell ratio (Fig. 5f). Conversely, the ratio of CD4^+^ to CD8^+^ T-cells was lower in respiratory samples (*p*=0.0065), indicating high prevalence of CD8^+^ T-cells in respiratory specimens (Fig. 5f).

Overall, neutrophils (CD16^+/-^) dominated in the respiratory samples of COVID-19 patients, with varying levels of monocytes/macrophages, T-cells (CD4^+^ and CD8^+^), NK cells, and B cells. T cells in the respiratory samples exhibited an activated and EM-like phenotype compared to paired blood samples, with lower CD4^+^ to CD8^+^ T-cell ratio.

### Respiratory antibody levels correlated with CD14^+^CD16^+^ monocytes, HLA-DR^+^ NK and CM/EM-like CD8^+^ T cells

To understand associations between immunological features in the respiratory specimens, correlations between plasma (cytokines, sIL-6R⍰, ADAMTS4, anti-RBD IgM, IgG, IgA) and cellular features were performed (Fig. 6a, b). Lower classical (CD14^+^CD16^-^) monocytes and higher intermediate (CD14^+^CD16^+^) monocytes correlated with higher FcγR2b SARS-CoV-2-Trimer-S-specific antibodies (*p*=0.0005; *p*=0.0004). Higher SARS-CoV-2-specific antibodies correlated with frequencies of (CM)-like (CD27^+^CD45RA^-^) and CD16^lo^neutrophils (*p*=0.0034; *p*=0.0076). Numbers of intermediate (CD14^+^CD16^+^) monocytes, HLA-DR^+^ NK, CD8^+^ T-cells (CM-like; EM-like) and CD16^hi^neutrophils correlated with antibody levels (*p*=0.0009-0.0039). Although cytokine profiles varied across COVID-19 respiratory samples, no significant differences were found in their cellular immunity (Fig. 6c). Unsupervised clustering also revealed distinct immunological features between respiratory and blood samples, with higher EM-like CD4^+^ and CD8^+^ T-cell frequencies and lower cell numbers (apart from neutrophils) in the respiratory samples (Fig. 6d).

**Fig. 6.**
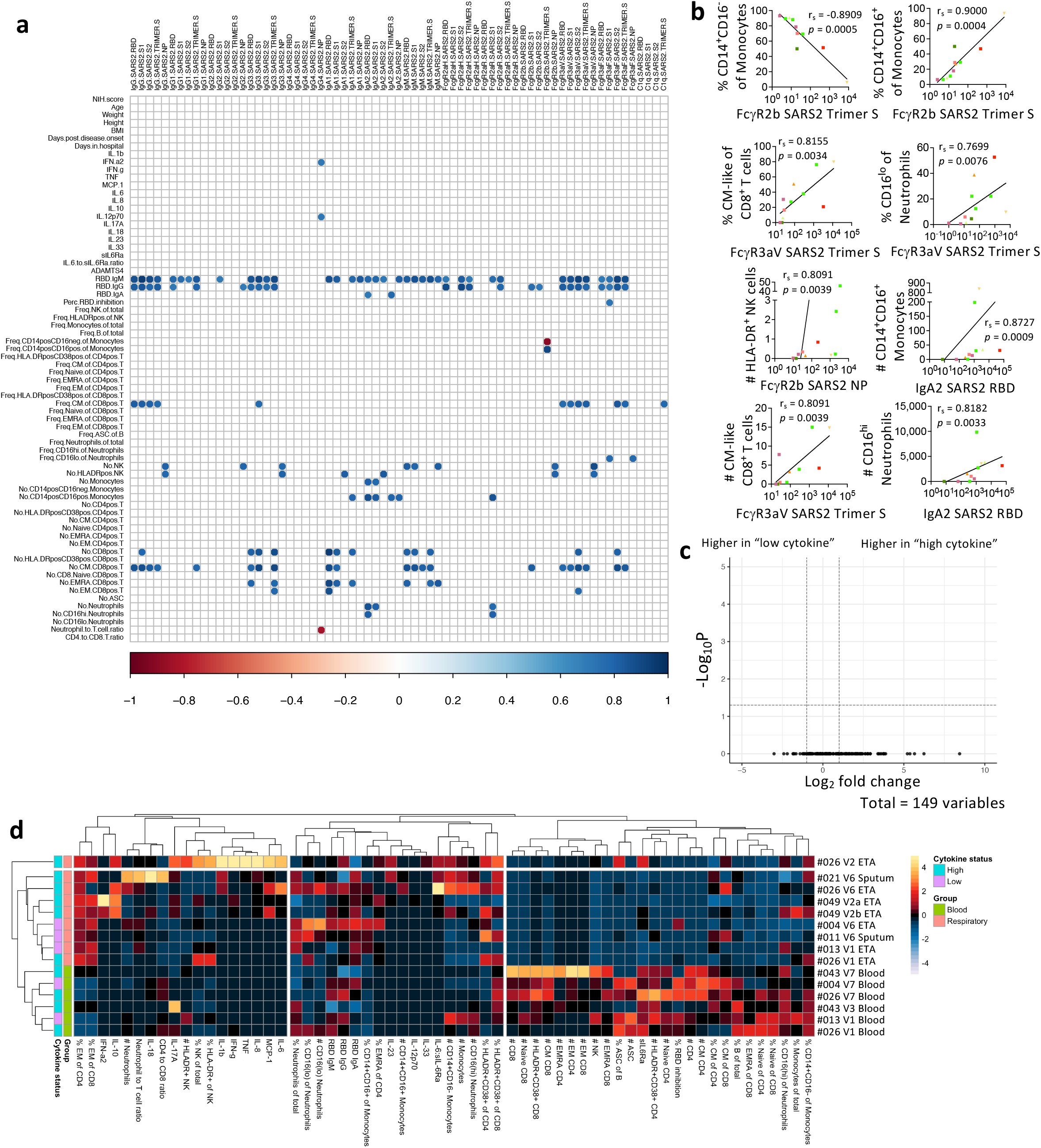
Airway SARS-CoV-2-specific antibody levels positively correlated with several immune cell types. **a**-**b** Correlation matrix and graphs in COVID-19 respiratory samples between multiplex and non-multiplex immune features. Correlation was determined with Spearman’s correlation and *p*-values of the correlation matrix were adjusted with False Discovery Rate adjustment. **c** Volcano plot showing fold difference of 149 cellular features in respiratory samples between patients with “low cytokine” (004, 011, 013) and “high cytokine” (016, 021, 026, 043, 049). **d** Heatmaps with unsupervised clustering of serological and cellular features in COVID-19 respiratory and blood samples.

### COVID-19 patients with higher NIH scores had more robust humoral immune responses

More patients with higher 4-5 NIH scores required ICU during hospitalization (Fig. 1d), while NIH scores of 2-3 were in the mild/moderate group. Although there were no differences in the overall cytokines/chemokines levels between the two NIH severity groups, IL-8 levels in the severe/critical group increased at discharge (V7) compared to admission (V1) (*p*=0.0004; Fig. 7a), indicating delayed or prolonged innate immune activation. Levels of sIL-6R⍰ were significantly higher in the severe/critical group than the mild/moderate group at both V1 (*p*=0.027) and V7 (*p*=0.0302), with the severe/critical group having higher sIL-6R⍰ and lower IL-6:sIL-6R⍰ ratio at V7 than V1 (Fig. 7a).

**Fig. 7.**
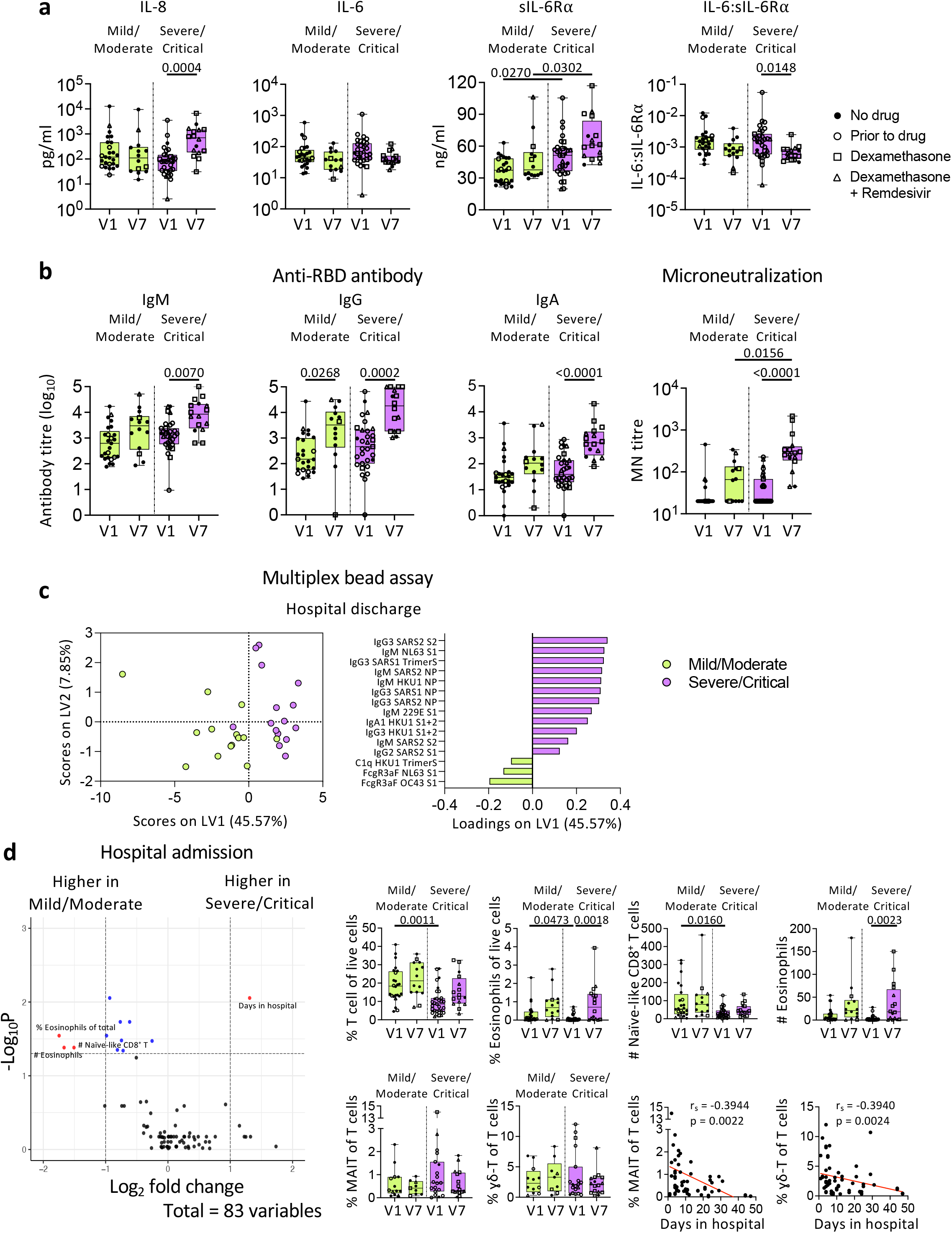
COVID-19 patients with higher NIH scores display more robust humoral immune responses towards SARS-CoV-2. **a** Levels of cytokines, sIL-6R⍰, and IL-6:sIL-6R⍰ ratio, **b** anti-RBD IgM, IgG, and IgA titres, microneutralization titres and **c** PLSDA scores and loadings plot of antibodies against human coronavirus between mild/moderate and severe/critical COVID-19 patients. **d** Volcano plot showing fold difference of 83 immunological features in blood samples between mild/moderate and severe/critical COVID-19 patients, and comparisons of cellular subset frequencies and correlation with days stayed in hospital. Statistical significance was determined with Kruskal-Wallis test followed by Dunn’s multiple comparisons test. Partial Least-Squares Discriminant Analysis was performed for antibodies measured with multiplex bead array assay. Volcano plots were created using a Wilcoxon rank-sum test and statistics were corrected with FDR adjustment. Correlation was determined with Spearman’s correlation. V1, hospital admission; V7, hospital discharge.

Anti-RBD IgG titres increased in both severity groups at discharge (*p*=0.0268; *p*=0.0002; Fig. 7b). The severe/critical group also displayed substantially higher microneutralization (MN) activity at discharge compared to admission (*p*<0.0001). PLSDA revealed that at discharge the severe/critical group had higher IgM and IgG antibodies targeting SARS-CoV-2 proteins compared to the mild/moderate group (Fig. 7c).

COVID-19 patients in the severe/critical group had comparable frequencies of immune cells, while they had lower T-cell and eosinophil frequencies (*p*=0.0011; *p*=0.0473) than the mild/moderate group at admission (V1; Fig. 7d). Interestingly, frequencies of mucosal associated invariant T (MAIT) cells and □δ T-cells negatively correlated with days stayed in hospital (*p*=0.0022 and *p*=0.0024 respectively; Fig. 7d).

Overall, while cytokine levels were similar between the two severity groups, patients with more severe symptoms had more robust antibody responses towards the SARS-CoV-2.

### Dexamethasone did not alter immune responses in COVID-19 patients

Effects of dexamethasone, corticosteroid anti-inflammatory drug, with/without remdesivir on immune responses are unclear. We found very few differences in immune profiles between patients with/without dexamethasone. IL-8 and sIL-6Rα levels at discharge were significantly higher than at admission in the dexamethasone (with/without remdesivir) group, but similar levels were observed without treatment (Fig. 8a). Patients on treatment had lower anti-inflammatory IL-10 levels at discharge (*p*=0.0281; Fig. 8a). Conversely, humoral responses of patients receiving drugs were not compromised. Patients receiving dexamethasone (with/without remdesivir) generated robust SARS-CoV-2-specific antibody responses (Fig. 8b). Given that 29/33 severe/critical patients were on treatment, compared to 7/27 mild/moderate patients, high antibody levels in the drug group were likely due to disease severity rather than drug treatment. PLSDA revealed that patients prior to drug therapy had higher antibodies against the NP of human coronavirus OC43 rather than SARS-CoV-2 (Fig. 8c). No significant differences were found in cellular responses, apart from lower T-cell frequency in the drug group (Fig. 8d).

**Fig. 8.**
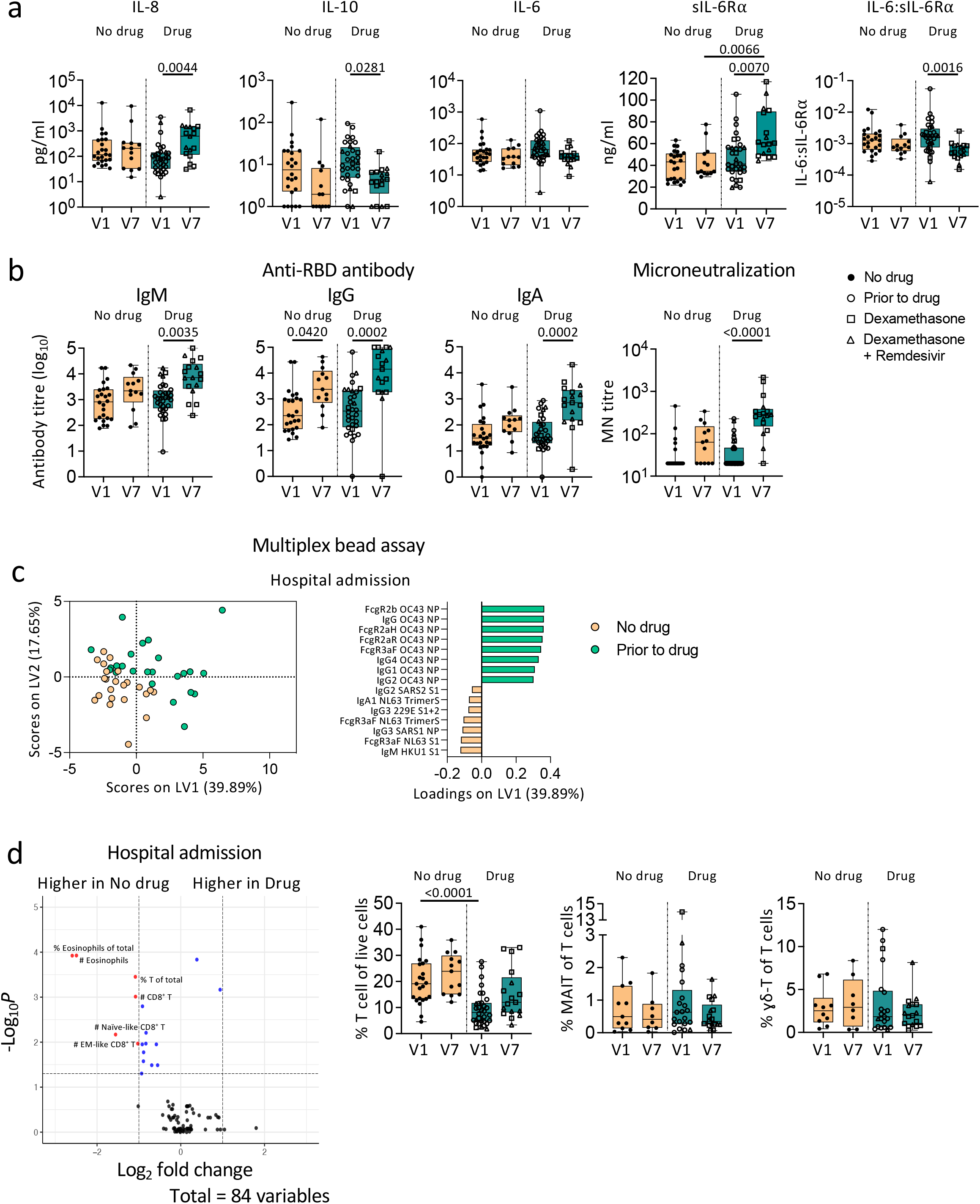
Dexamethasone treatment does not alter humoral immune responses towards SARS-CoV-2 in COVID-19 patients. **a** Levels of cytokines, sIL-6R⍰, and IL-6:sIL-6R⍰ ratio, **b** anti-RBD IgM, IgG, and IgA titres, microneutralization titres and **c** antibodies against human coronavirus **d** cellular immune subset frequencies between COVID-19 patients with or without dexamethasone treatment (with/without remdesivir). Statistical significance was determined with Kruskal-Wallis test followed by Dunn’s multiple comparisons test. Partial Least-Squares Discriminant Analysis was performed for antibodies measured with multiplex bead array assay. Volcano plots were created using a Wilcoxon rank-sum test and statistics were corrected with FDR adjustment. V1, hospital admission; V7, hospital discharge.

## DISCUSSION

Immunity to SARS-CoV-2 in the respiratory tract, the primary site of infection, is incompletely understood. We found discordant inflammatory status in the respiratory tract of COVID-19 patients, whereas non-COVID-19 hospitalized patients had consistently high respiratory cytokine levels. While high SARS-CoV-2-specific IgG and IgM were detected in COVID-19 respiratory samples, IgG with FcγR binding profiles were more prominent in blood. We found higher frequencies of neutrophils, intermediate CD14^+^CD16^+^monocytes, activated HLA-DR^+^CD38^+^, EM-like CD4^+^ and CD8^+^ T-cells in COVID-19 respiratory compared to blood samples, and similar immune responses with dexamethasone (with/without remdesivir) treatment.

High levels of cytokines are commonly found in blood of COVID-19 patients^29, 30, 31, 32^. In respiratory samples, variable cytokine levels (IL-10, IL-17A, IL-18) were detected, while monocyte chemoattractants (MCP-1, MIP-1α, MIP-1β) and innate cytokines (IL-6, IL-10) were at high levels^18, 19^. We found hypercytokinemia in respiratory samples compared to blood, but only in selected COVID-19 patients with high IL-6, IL-8, and MCP-1, indicating an inflammatory environment that attracts leukocytes, including neutrophils and monocytes^33,34^. Since most patients did not have similar cytokine profiles in blood and respiratory samples, thus measuring both blood and respiratory inflammation might be needed to accurately determine the inflammatory status of the patients.

SARS-CoV-2-specific IgG and IgA were detected previously in BALF, sputum and saliva of COVID-19 patients^35, 36, 37^. We found detectable anti-RBD IgM, IgA and IgG in COVID-19 respiratory samples, with higher IgM and IgG than non-COVID-19 respiratory samples. Although only two COVID-19 ETAs had detectable neutralizing activity, both samples had high levels of anti-RBD IgM, IgG, and IgA. While most attention focused on IgA at mucosal surfaces, SARS-CoV-2-specific IgM also was detected in sputum, BALF and saliva from severely-ill COVID-19 patients^35, 37^. Unlike IgAs^38^, anti-RBD IgM in saliva of COVID-19 patients strongly correlated with serum levels^37^. As IgM might affect the immunopathology in the respiratory tract, this warrants further investigations.

Similar to previous studies, neutrophils dominated in COVID-19 respiratory samples^21^. Longitudinal ETA samples indicated increases in cellular infiltrates during disease progression, while presence of CD16^lo^ neutrophils showed recruitment of immature neutrophils likely derived from emergency myelopoiesis in bone marrow^39^. RNA-sequencing of COVID-19 BALF neutrophils found similar immature states^40^. Higher frequencies of intermediate CD14^+^CD16^+^ monocytes and activated CD38^+^HLA-DR^+^CD4^+^ T-cells in respiratory samples reveal activated signatures in the respiratory tract. Although low in frequency, high activated respiratory T-cells are associated with improved survival in COVID-19^18^.

As an anti-inflammatory drug, dexamethasone can reduce proinflammatory cytokines including IL-6 and IL-8^41, 42^. We found no significant difference in cytokine/chemokine levels between COVID-19 patients receiving dexamethasone and untreated patients. Although it has been speculated that dexamethasone can reduce the ability of antibody production in B cells^43^, we showed similar antibody levels in patients receiving dexamethasone. Therefore, severely-ill COVID-19 patients might benefit from dexamethasone treatment as reported^5, 6, 7^, and such treatment does not dampen humoral immunity.

There are limitations to the current study. Firstly, ETA samples were only collected from patients with severe disease requiring invasive oxygen support, therefore, it is unclear whether COVID-19 patients with milder symptoms had less robust immune responses in the respiratory tract. Additionally, most patients in the severe/critical group received dexamethasone, which could be an intercorrelating factor for the differences observed between severity groups. Moreover, while the non-COVID-19 controls provided insights onto the immune status in hospitalized individuals, the comparisons will benefit more if there were larger numbers of non-COVID patients with more homogenous diseases.

Overall, innate and adaptive immune responses are generated in respiratory and blood samples of COVID-19 patients. While immunological features detected in the peripheral blood can be associated with robust immune responses and predict clinical outcomes, monitoring immune responses in the respiratory samples can be of a benefit prior to initiation of therapeutic interventions for COVID-19 patients.

## Data Availability

Raw data are available upon request.

## ACKNOWLEDGMENTS

We acknowledge all DRASTIC (The use of cytokines as a preDictoR of disease Severity in criTically Ill COVID-19) investigators from Austin Health, and thank the participants involved. We acknowledge Francesca L. Mordant and Kanta Subbarao for their contributions to the microneutralization assay. We acknowledge Adam K. Wheatley for kindly providing SARS-CoV-2 and HKU-1 spike trimers and Bruce D. Wines and P. Mark Hogarth for kindly providing FcγR dimers. The RBD proteins were produced under HHSN272201400008C and obtained through BEI Resources, NIAID, NIH: Spike Glycoprotein RBD from SARS-Related Coronavirus 2, Wuhan-Hu-1 with C-Terminal Histidine Tag, Recombinant from HEK293F Cells, NR-52366. We thank the staff at the diagnostic microbiology laboratories at Austin Pathology, Melbourne Pathology, Dorevitch Pathology, 4Cyte Pathology, Microbiological Diagnostic Unit Public Health Laboratory, Northern Pathology Victoria, Eastern Health Pathology for performing initial diagnostic testing for the detection of SARS-CoV-2 nucleic acid. This work was supported by the NHMRC Leadership Investigator Grant to KK (#1173871), Research Grants Council of the Hong Kong Special Administrative Region, China (#T11-712/19-N) to KK, the MRFF Award (#1202445) to KK and AWC, NIH contract CIVC-HRP (HHS-NIH-NIAID-BAA2018) to PGT and KK, NHMRC Senior Principal Research Fellowship (#1117766) to DIG, NHMRC Emerging Leadership Level 1 Investigator Grant to THON (#1194036) and NHMRC Early Career Fellowships to HFK (#1160333), CLG (#1160963) and JAT (#1139902). WZ and JRH are supported by the Melbourne Research Scholarship from The University of Melbourne. LH is supported by the Melbourne International Research Scholarship (MIRS) and the Melbourne International Fee Remission Scholarship (MIFRS) from The University of Melbourne. We acknowledge the Melbourne Cytometry Platform (Peter Doherty Institute and Melbourne Brain Centre nodes) for provision of flow cytometry services. PGT is supported by NIH NIAID R01 AI136514-03 and ALSAC at St. Jude.

## AUTHOR CONTRIBUTIONS

KK, THON, CLG and JAT supervised the study. KK, THON, CLG, JAT, WZ, BYC, KJS, LK, HFK, SN and AWC designed the experiments. WZ, BYC, KJS, LK, ERH, LH, LCR, JRH, LFA, HFK, JAN, MG, SN and THON performed and analysed experiments. WZ, TMA, SKS, CYL, PA, TS, NLS, and KBA analysed data. DFB, FA, FK and PGT provided reagents. FJ, EM, JK, KYLC, GD, AC, JED, NEH, OCS, JAT, CLG recruited the patient cohorts and provided clinical data. WZ, SLL, LMW, NJCK, DIG, LKM, PGT, SN, KBA, AWC, JAT, CLG, THON and KK provided intellectual input into the study design and data interpretation. WZ, THON and KK wrote the manuscript. All authors reviewed and approved the manuscript.

## METHODS

### DRASTIC study participants and specimens

We enrolled 60 SARS-CoV-2 PCR-positive patients admitted to Austin Health (Victoria, Australia) and six PCR-negative patients as negative serological controls. Two COVID-19 patients and three SARS-CoV-2 PCR-negative patients died during the study. Peripheral blood was collected in heparinized or ethylenediaminetetraacetic acid (EDTA) tubes during hospitalization. Peripheral blood monocular cells (PBMCs) were isolated via Ficoll-Paque separation. Single cell suspensions were isolated from tissues as previously described^44, 45^. ETA samples were obtained as part of routine suctioning of the endotracheal tube airway and involved the passage of a catheter for suctioning into a sterile sputum trap. Sputum samples were spontaneously collected into a sterile container. Pleural fluid was collected by thoracentesis as part of a routine procedure. The thoracentesis involved percutaneous insertion of a catheter into the pleural space and collection of pleural fluid into a sterile container. Demographic, clinical and sampling information for COVID-19 patients are described in Supplementary Table 1.

### Ethics statement

Experiments conformed to the Declaration of Helsinki Principles and the Australian National Health and Medical Research Council Code of Practice. Written informed consent was obtained from all blood donors prior to the study. The study was approved by the Austin Health (HREC/63201/Austin-2020) and the University of Melbourne (#2057366.1, #2056901.1 and #1955465.3) Human Research Ethics Committees.

### Genomic sequencing and bioinformatic analysis

Extracted RNA from RT-PCR positive samples underwent tiled amplicon PCR and Illumina short-read sequencing, quality control, consensus sequence generation and alignment as previously described^46^. A single sequence per patient was used for phylogenetic analysis^22^, with a maximum-likelihood phylogenetic tree generated using IQ-Tree^47^ and visualized using the *ggtree* package in R^48^. Genomic clusters were defined using a hierarchical clustering algorithm; genomic transmission networks grouped multiple clusters supported by epidemiological and genomic data.

### Phenotypic whole blood immune analyses

Fresh whole blood (200μl per stain) was used to measure CD4^+^CXCR5^+^ICOS^+^PD1^+^ follicular T cells (Tfh) and CD3^-^CD19^+^CD27^hi^CD38^hi^ antibody-secreting B cell (ASC; plasmablast) populations as described^15, 49^ as well as activated HLA-DR^+^CD38^+^CD8^+^ and HLA-DR^+^CD38^+^CD4^+^ T cells, intermediate CD14^+^CD16^+^ and classical CD14^+^ monocytes, activated CD3^-^CD56^+^ NK cells, MAIT cells, [δ-T cells, as per the specific antibody panels (Supplementary Table 6; gating strategy is presented in Supplementary Fig. 4b, c). After whole blood was stained for 20 minutes at room temperature in the dark, samples were lysed with BD FACS Lysing solution, washed and fixed with 1% PFA. AccuCheck Counting Beads (Thermo Fisher Scientific) were added for calculating absolute numbers just prior to acquisition. All samples were acquired on a LSRII Fortessa (BD). Flow cytometry data were analyzed using FlowJo v10 software.

### Phenotypic immune analyses in respiratory samples

Respiratory samples (ETA, sputum, or pleural fluid) were diluted in PBS and centrifuged. Supernatant was collected and the pellet was filtered through a 45μm filter prior to separation of respiratory fluid and cellular contents. The respiratory fluid was frozen at -20°C, and the cell pellet was washed with EDTA-BSS. Washed cells were stained with FcR block (Miltenyi Biotec) for 15 minutes followed by 30 minutes staining on ice with specific antibody panels (Supplementary Table 6). After fixing with 1% PFA, the samples were acquired on a LSRII Fortessa (BD). AccuCheck Counting Beads were added for calculating absolute numbers just prior to acquisition. Flow cytometry data were analyzed using FlowJo v10 software.

### SARS-CoV-2 RBD ELISA

RBD-specific ELISA for detection of IgM, IgG and IgA antibodies was performed as previously described^31, 50, 51^, using flat bottom Nunc MaxiSorp 96-well plates (Thermo Fisher Scientific) for antigen coating (2µg/ml), blocking with PBS (with w/v 1% BSA) and serial dilutions in PBS (with v/v 0.05% Tween and w/v 0.5% BSA). Inter- and intra-experimental measurements were normalised using a positive control plasma from a COVID-19 patient run on each plate. Endpoint titres were determined by interpolation from a sigmodial curve fit (all R-squared values >0.95; GraphPad Prism 9) as the reciprocal dilution of plasma that produced >15% (for IgA and IgG) or >30% (for IgM) absorbance of the positive control at a 1:31.6 (IgG and IgM) or 1:10 dilution (IgA). Seroconversion was defined when titres were above the mean titre (plus 2 standard deviations) of non-COVID-19 control respiratory or plasma samples.

### Microneutralization assay

Microneutralization activity of serum samples was assessed as previously described^52^. SARS-CoV-2 isolate CoV/Australia/VIC01/2020^53^ was propagated in Vero cells and stored at -80°C. Sera were heat-inactivated at 56°C for 30 min and serially diluted. Residual virus infectivity in the serum/virus mixtures was assessed in quadruplicate wells of Vero cells incubated in serum-free media containing 1μg/ml of TPCK trypsin at 37°C and 5% CO_2_. Viral cytopathic effect was read on day 5. The neutralizing antibody titer was calculated using the Reed-Muench method^52^.

### SARS-CoV-2 surrogate virus neutralisation test (sVNT)

The plasma samples were tested in neat, and the respiratory samples were tested at 1:9 dilution or at their original dilutions for more diluted samples. The sVNT blocking ELISA assay (manufactured by GenScript, NJ, USA) was carried out essentially as described^51^, which detects circulating neutralizing SARS-CoV-2 antibodies that block the interaction between RBD and ACE2 on the cell surface receptor of the host. HRP-conjugated recombinant SARS-CoV-2 RBD fragment bound to any circulating neutralizing antibodies to RBD preventing capture by the human ACE2 protein in the well, which was subsequently removed in the following wash step. Substrate reaction incubation time was 20 mins at room temperature and results were read spectrophotometrically. Colour intensity was inversely dependent on the titre of anti-SARS-CoV-2 neutralizing antibodies.

### Coupling of carboxylated beads

As previously described^25^, a custom multiplex bead array was designed and coupled with SARS-CoV-2 spike 1 (Sino Biological), spike 2 (ACRO Biosystems), RBD (BEI Resources) and nucleoprotein (ACRO Biosystems), as well as SARS and hCoV (229E, NL63, HKU1, OC43) spikes and nucleoproteins (Sino Biological) (Supplementary Fig. 5). In addition, SARS-CoV-2 and HKU-1 spike trimers (kind gifts from Adam K. Wheatley), as well as SARS-CoV and NL63 spike trimers (BPS Bioscience) were also coupled. Tetanus toxoid (Sigma-Aldrich), influenza hemagglutinin (H1Cal2009; Sino Biological) and SIV gp120 (Sino Biological) were also included in the assay as positive and negative controls respectively. Antigens were covalently coupled to magnetic carboxylated beads (Bio Rad) using a two-step carbodiimide reaction and blocked with 0.1% BSA, before being resuspended and stored in PBS 0.05% sodium azide.

### Luminex bead-based multiplex assay

Using the coupled beads mentioned above, a custom CoV multiplex assay was formed to investigate the isotypes and subclasses of pathogen-specific antibodies present in collected plasma samples^25^. Briefly, 20µl of working bead mixture (1000 beads per bead region) and 20µl of diluted plasma (final dilution 1:200) or 20µl of diluted respiratory secretions (final dilution 1:800) were added per well and incubated overnight at 4°C on a shaker. Fourteen different detectors were used to assess pathogen-specific antibodies. Single-step detection was done using phycoerythrin (PE)- conjugated mouse anti-human pan-IgG, IgG1-4 and IgA1-2 (Southern Biotech; 1.3µg/ml, 25µl/well). C1q protein (MP Biomedicals) was first biotinylated (Thermo Fisher Scientific), then tetramerized with Streptavidin R-PE (SAPE; Thermo Fisher Scientific) before dimers or tetrameric C1q-PE were used for single-step detection. For the detection of FcγR-binding, soluble recombinant FcγR dimers (higher affinity polymorphisms FcγRIIa-H131, lower affinity polymorphisms FcγRIIa-R131, FcγRIIb, higher affinity polymorphisms FcγRIIIa-V158, lower affinity polymorphisms FcγRIIIa-F158; 1.3µg/ml, 25µl/well; kind gifts from Bruce D. Wines and P. Mark Hogarth) were first added to the beads, washed, and followed by the addition of SAPE. For the detection of IgM, biotinylated mouse anti-human IgM (mab MT22; Mabtech; 1.3µg/ml, 25µl/well) was first added to beads, washed, followed by SAPE. Assays were read on the Flexmap 3D and performed in duplicates.

### Data normalization

For all multivariate analysis, Tetanus, H1Cal2009, and BSA antigens (positive controls) were removed, as well as SIV (negative control). Low signal features were removed when the 75^th^ percentile response for the feature was lower than the 75^th^ percentile of the BSA positive control. Right shifting was performed on each feature (detector–antigen pair) individually if it contained any negative values, by adding the minimum value for that feature back to all samples within that feature. Following this, all data were log-transformed using the following equation, where x is the right-shifted data and y is the right-shifted log-transformed data: y = log10(x + 1). This process transformed the majority of the features to having a normal distribution. In all the subsequent multivariate analyses, the data were furthered normalized by mean centering and variance scaling each feature using the z-score function in Matlab. Plasma and respiratory samples were analysed separately. When analysing samples at time of hospital discharge, to adjust for the confounder of time from symptom onset, each of the features were iteratively regressed with ordinary least squares regression, using the residuals as input for the analysis^54^.

### Feature selection using Elastic Net/PLSDA

To determine the minimal set of features (signatures) needed to predict categorical outcomes (COVID-19 diagnosis, NIH scores, drug therapies), a three-step process was developed^55^. First, the data were randomly sampled without replacement to generate 2000 subsets. The resampled subsets spanned 80% of the original sample size, or sampled all classes at the size of the smallest class for categorical outcomes, which corrected for any potential effects of class size imbalances during regularization. Elastic-Net regularization was then applied to each of the 2000 resampled subsets to reduce and select features most associated with the outcome variables. The Elastic-Net hyperparameter, α, was set to have equal weights between the L1 norm and L2 norm associated with the penalty function for least absolute shrinkage and selection (LASSO) and ridge regression, respectively ^56^. By using both penalties, Elastic-Net provides sparsity and promotes group selection. The frequency at which each feature was selected across the 2000 iterations was used to determine the signatures by using a sequential step-forward algorithm that iteratively added a single feature into the PLSDA model starting with the feature that had the highest frequency of selection, to the lowest frequency of selection. Model prediction performance was assessed at each step and evaluated by 10-fold cross-validation classification error for categorical outcomes. The model with the lowest classification error within a 0.01 difference between the minimum classification error was selected as the minimum signature. If multiple models fell within this range, the one with the least number of features was selected and if there was a large disparity between calibration and cross-validation error (over-fitting), the model with the least disparity and best performance was selected.

### PLSDA

Partial least squares discriminant analysis (PLSDA), performed in Eigenvectors PLS toolbox in Matlab, was used in conjunction with Elastic-Net, described above, to identify and visualize signatures that distinguish categorical outcomes (COVID-19 diagnosis, NIH scores, drug therapies). This supervised method assigns a loading to each feature within a given signature and identifies the linear combination of loadings (a latent variable, LV) that best separates the categorical groups. A feature with a high loading magnitude indicates greater importance for separating the groups from one another. Each sample is then scored and plotted using their individual response measurements expressed through the LVs. The scores and loadings can then be cross-referenced to determine which features are loaded in association with which categorical groups (positively loaded features are higher in positively scoring groups, etc.). All models go through 10-fold cross-validation, where iteratively 10% of the data is left out as the test set, and the rest is used to train the model. Model performance is measured through calibration error (average error in the training set) as well as cross-validation error (average error in the test set), with values near 0 being best. All models were orthogonalized to enable clear visualization of results.

### Hierarchical clustering

We visualized the clustering of DRASTIC respiratory and blood samples based on only SARS-CoV-2 antigens or all features using unsupervised average linkage hierarchical clustering of normalized data using MATLAB 2017b (MathWorks, Natick, MA). Euclidean distance was used as the distance metric.

### Cytokine analysis

Patients’ plasma and respiratory samples were measured for IL-1β, IFN-α2, IFN-γ, TNF, MCP-1 (CCL2), IL-6, IL-8 (CXCL8), IL-10, IL-12p70, IL-17A, IL-18, IL-23 and IL-33 using the LEGENDplex™ Human Inflammation Panel 1 kit, as per manufacturer’s instructions (BioLegend, San Diego, CA, USA).

### sIL-6Rα and ADAMTS4 ELISAs

Soluble protein levels were all measured using DuoSet ELISA kits for each protein (R&D Systems, Minneapolis, MN, USA) according to the manufacturer’s instructions. DuoSet ELISA ancillary reagent kit (R&D Systems) was used for respiratory fluids and in-house reagents with the same composition were used for plasma samples. In brief, 96-well R&D ELISA microplates (respiratory fluids) or 96-well Nunc Maxisorp ELISA plates (ThermoFisher, plasma) were coated with capture antibody overnight, followed by blocking with 1% w/v BSA for a minimum of 1 hour. Samples and standard proteins were added and incubated for 2 hours at room temperature, followed by detection antibody for a further 2 hours. Lastly, streptavidin-HRP, substrate solution and stop solution (2N H_2_SO_4_) were added subsequently for 20 minutes each. Plasma samples were diluted in 1:300 for sIL-6Rα ELISAs. Respiratory fluids were diluted in 1:50/1:150 for sIL-6Rα ELISA accounting in the original dilution factors and tested without further dilution for ADAMTS4 ELISA.

### Computational flow cytometry analysis

Computational analysis of data was performed using the Spectre R package^26^ (https://github.com/ImmuneDynamics/Spectre). Samples were initially prepared in FlowJo, and populations of interest were exported as CSV files containing raw (scale value) data. In R, data were subject to arcsinh transformation and clustering using FlowSOM ^27^. For visualisation, cells in the myeloid panel were subjected to sample-weighted downsampling based on absolute cells/uL counts in the blood, whereas cells in the lymphoid panel were unchanged to preserve samples with low cell numbers. Cells from the lymphoid panel, and downsampled cells from the myeloid panel, were then distributed in 2D via dimensionality reduction (DR) using Fast Interpolation-based t-distributed Stochastic Neighbour Embedding (FIt-SNE)^28^. Data from both the lymphoid and myeloid panel were subject to two rounds of clustering and DR. The initial round of clustering and DR was used to filter out cellular debris and non-immune cells exhibiting high autofluorescence, using arcsinh transformed expression of CD45, CD3, CD4, CD8, CD14, CD16, CD19, CD64, CD66b, CD38, HLA-DR, and Live-Dead for the myeloid panel; and CD45, CD3, CD4, CD8, TCR-γδ, CD45RA, CD27, CD56, CD19, CD16, CD14, CD38, HLA-DR, PD-1, and Live-Dead for the lymphoid panel. A second round of clustering and DR was then used for detailed immunophenotyping of cells in the respiratory tract, using arcsinh transformed expression of CD4, CD8, CD14, CD16, CD38, CD45, CD64, CD66b, HLA-DR, and Live-Dead for the myeloid panel; and CD45, CD3, CD4, CD8, TCR-γδ, CD45RA, CD27, CD56, CD19, CD16, CD38, and HLA-DR for the lymphoid panel.

Immune cell lineages were manually annotated based on marker expression: neutrophils (FSC^int^SSC^int^CD66b^+^), monocytes (FSC^int^CD14^+^), B cells (CD19^+^), NK cells (CD56^+^), gamma-delta T cells (TCR-γδ^+^), CD4^+^ T cells (CD4^+^CD14^-^), CD8^+^ T cells (CD8^+^), and dead cells (Live-Dead^+^). Interestingly, CD3 expression on T cell subsets was not apparent in respiratory samples, and we did not find distinct eosinophil phenotypes. Subsequently, population subsets were manually annotated based on marker expression: neutrophils (CD16^hi^, CD16^lo^, CD16^-^, dead neutrophils Live-Dead^+^), monocytes (classical CD14^+^CD16^-^ and intermediate CD14^+^CD16^+^), B cells (naïve CD27^-^CD38^-^, memory CD27^+^CD38^-^, ASC CD27^+^CD38^+^), NK cells (CD56^bri^ and CD56^dim^), CD4^+^ T cells (naïve-like CD45RA^+^CD27^+^, EMRA-like CD45RA^+^CD27^-^, CM-like CD45RA^-^CD27^+^, and EM-like CD45RA^-^CD27^-^), and CD8^+^ T cells (naïve-like CD45RA^+^CD27^+^, EMRA-like CD45RA^+^CD27^-^, CM-like CD45RA^-^CD27^+^, and EM-like CD45RA^-^CD27^-^). Given that NK cells were classified as CD56^+^ cells, the subset might include other unconventional T cells. Subsets were evaluated for expression of CD38, HLA-DR, and PD-1 expression using manual gating in FlowJo.

Volcano plots and heatmaps were created using the Spectre R package^26^, where comparisons were performed using a Wilcoxon rank-sum test (equivalent to the Mann-Whitney test) with the wilcox.test function in R. Statistics displayed in volcano plots were corrected with a False Discovery Rate (FDR) adjustment.

### Statistical analyses

Statistical significance was assessed using Mann-Whitney, Wilcoxon signed-rank test or Kruskal-Wallis test with Dunn’s correction for multiple comparisons in Prism 9 (GraphPad) unless stated otherwise. Correlations were assessed using Spearman’s correlation coefficient (*r*_s_) and visualized in R 3.6.2 as heatmaps using the corrplot package or using the online Morpheus heatmap software (https://software.broadinstitute.org/morpheus; the Broad Institute, MA, USA) and p-values of correlations were corrected for multiple comparisons by FDR in R 3.6.2. *P-*values lower than 0.05 were considered statistically significant.

## Data availability

The published article includes all datasets generated or analyzed during the study.

