## Supplementary Figures for "Immune responses in COVID-19 respiratory tract and blood reveal mechanisms of disease severity"

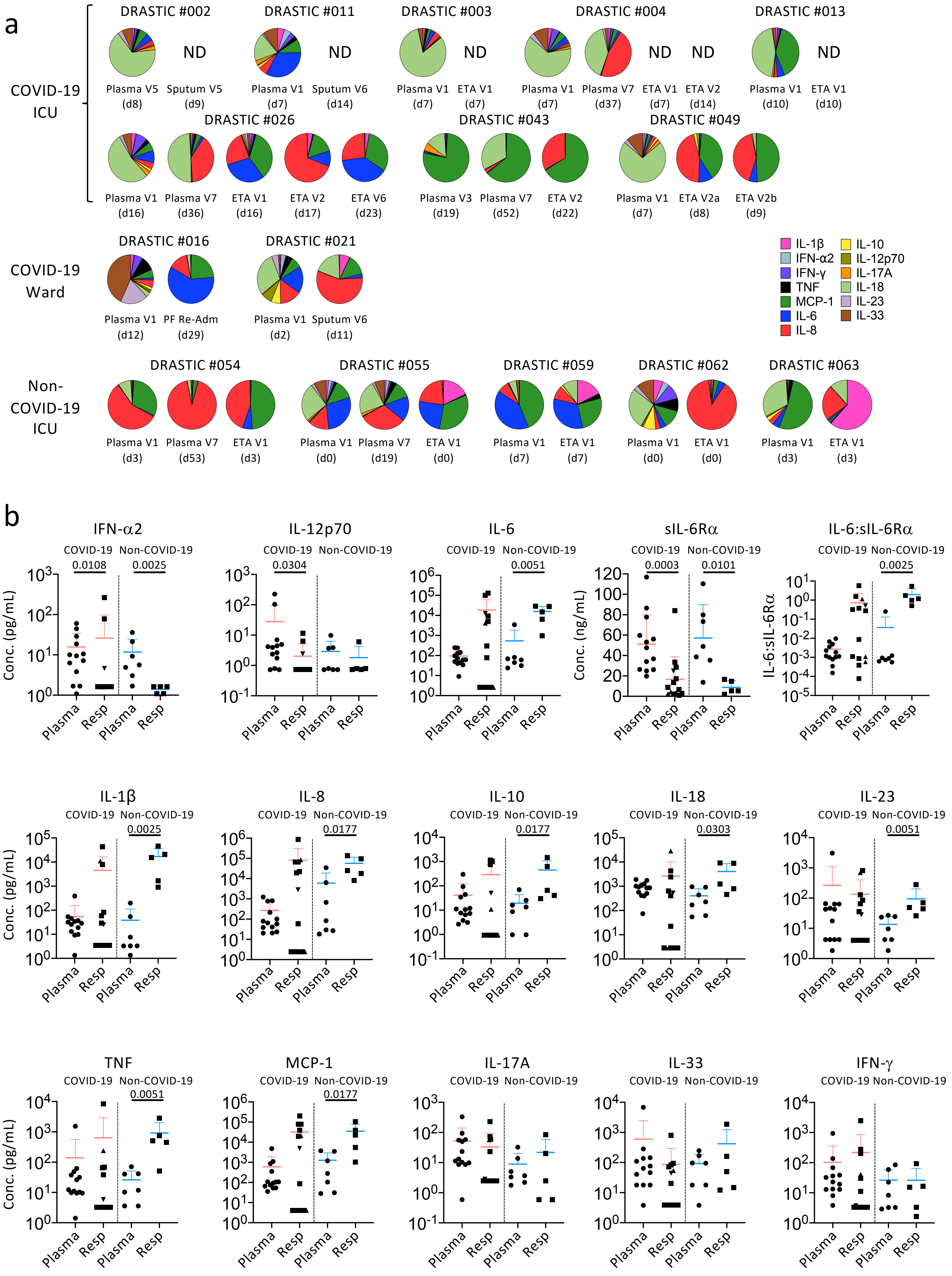


Supplementary Fig. 1 Zhang *et al*

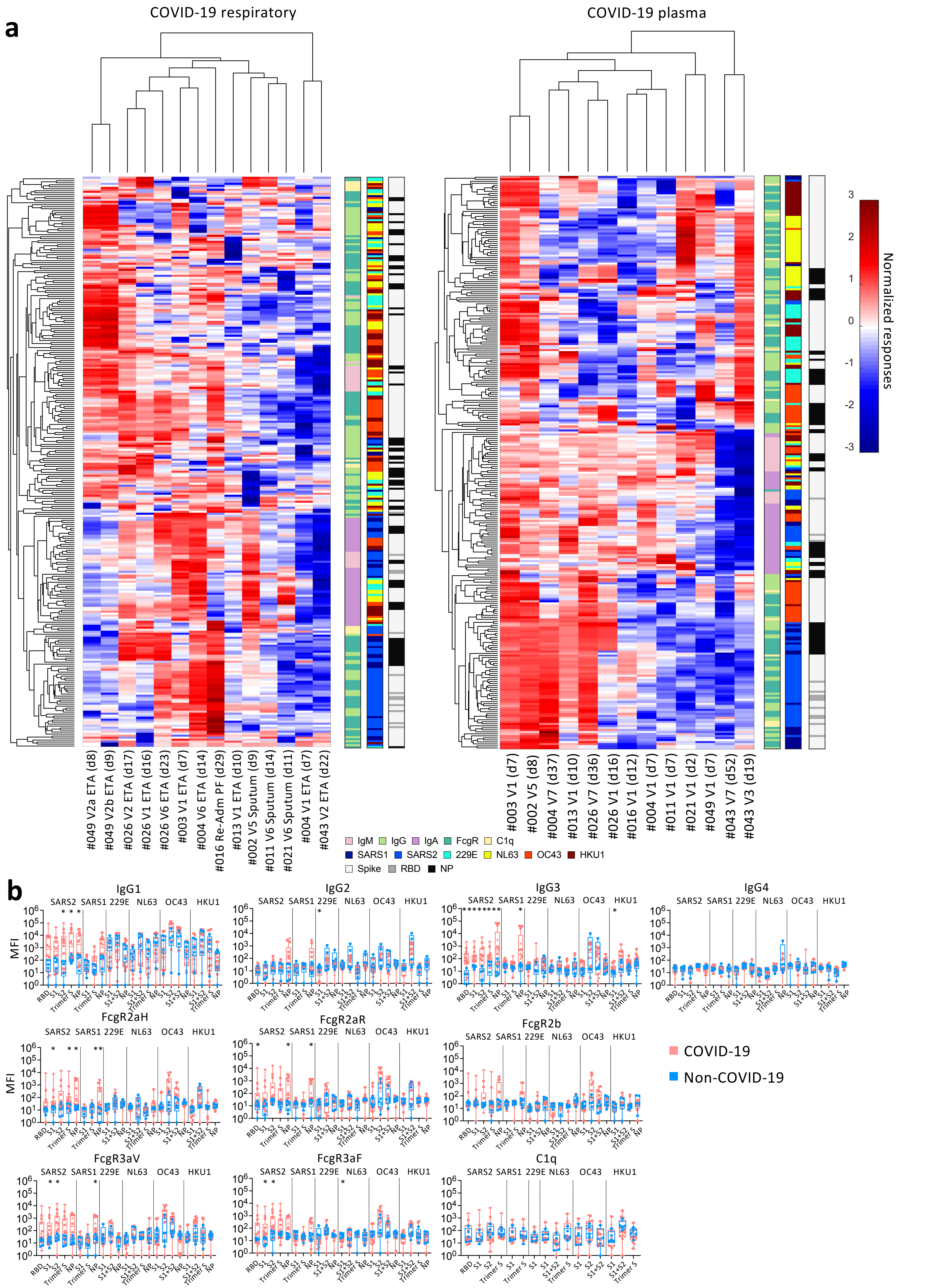


Supplementary Fig. 2 Zhang *et al*

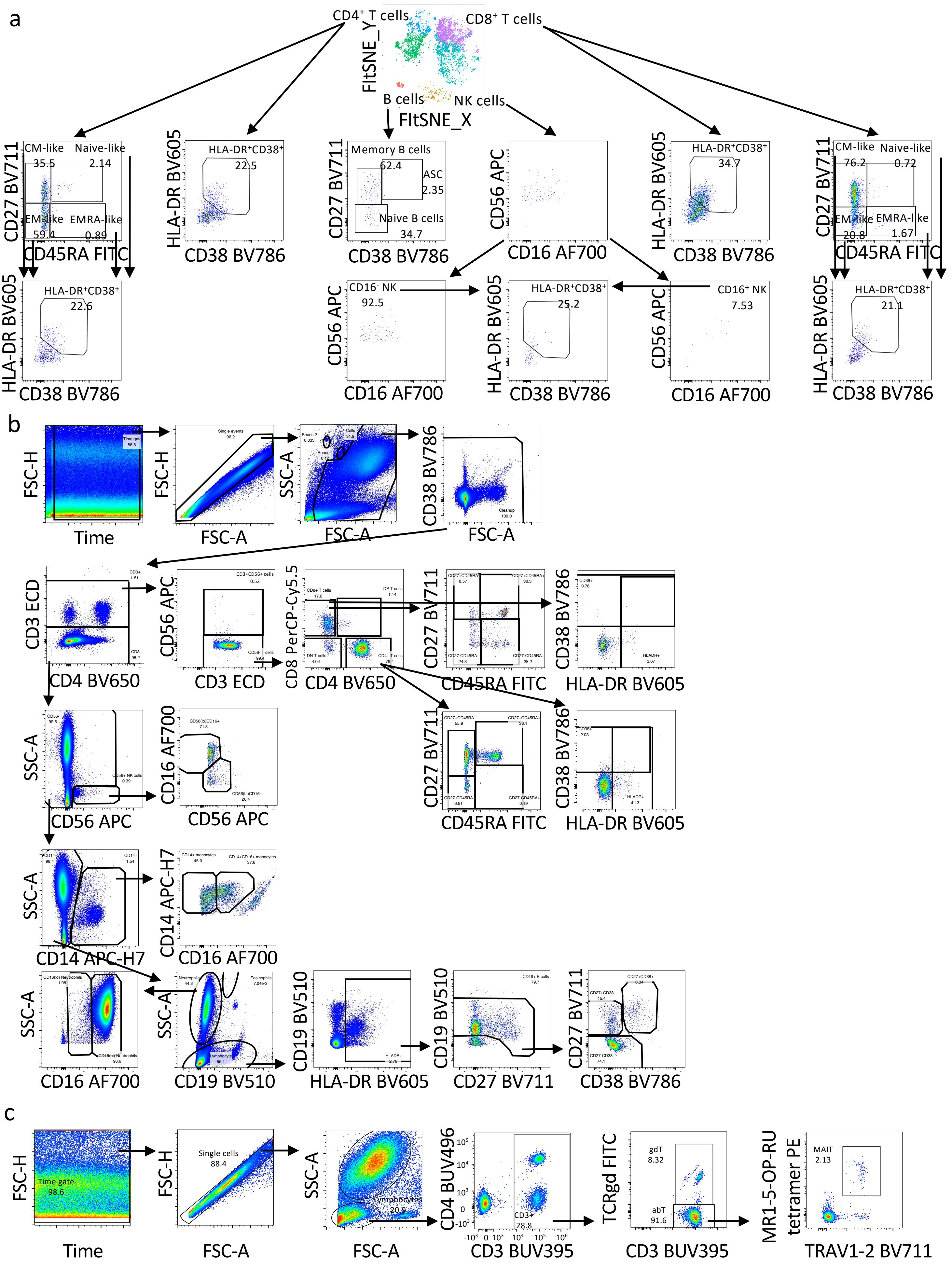


Supplementary Fig. 3 Zhang *et al*

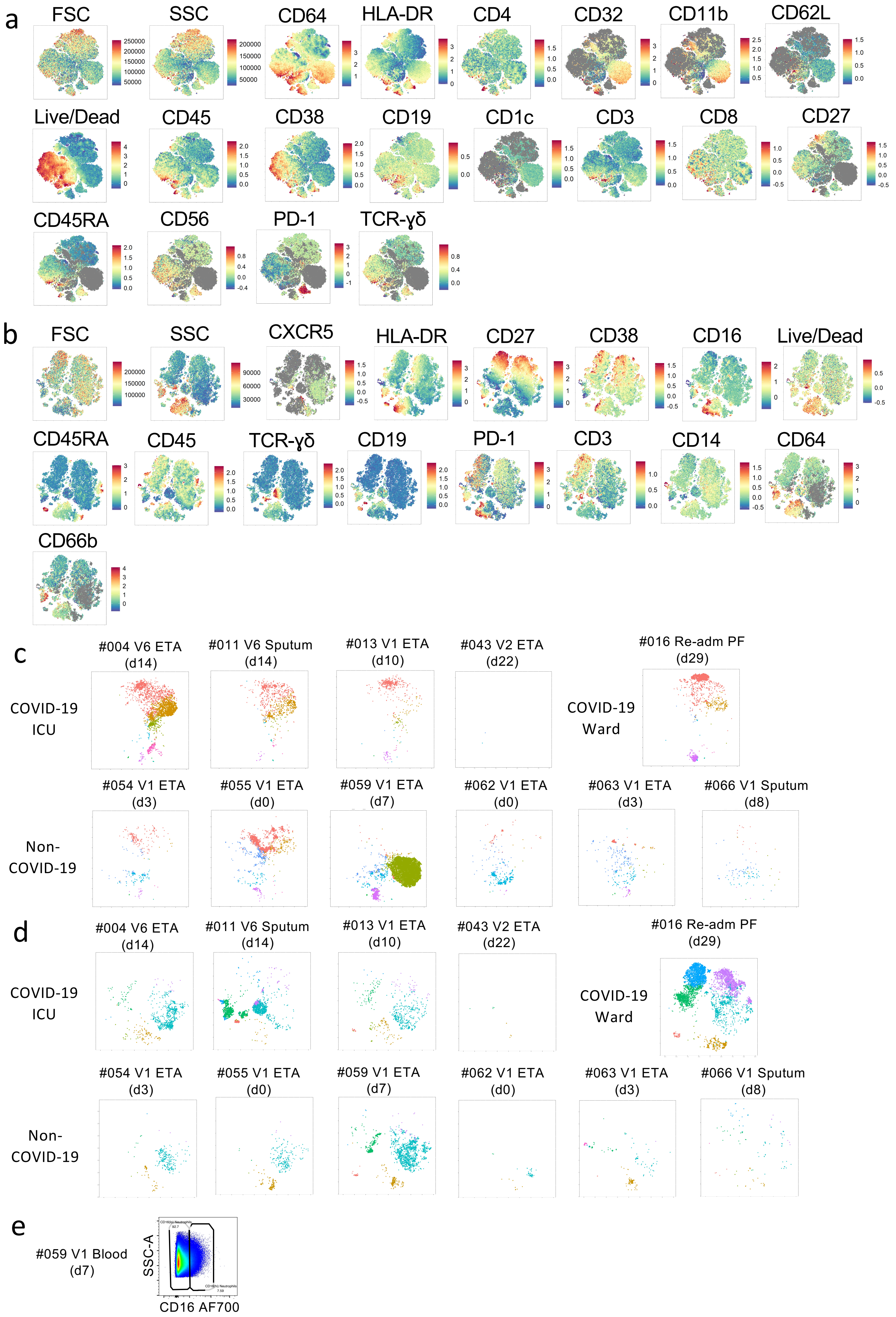


Supplementary Fig. 4 Zhang *et al*
