## Supplementary Information for "Immune responses in COVID-19 respiratory tract and blood reveal mechanisms of disease severity"

**Supplementary Fig. 1 High cytokine levels in non-COVID-19 respiratory samples. a** Distribution of 13 cytokines and chemokines (IL-1β, IFN-⍺2, IFN-γ, TNF, MCP-1, IL-6, IL-8, IL-10, IL-12p70, IL-17A, IL-18, IL-23, and IL-33) in each individual that respiratory and paired blood samples were collected. **b** Comparison of levels of cytokines, chemokines, sIL-6R⍺, and IL-6:sIL-6R⍺ ratio between plasma and respiratory samples for COVID-19 and non-COVID-19 patients. Lines indicate average+SD. Statistical significance was determined with Mann-Whitney test.

**Supplementary Fig. 2 Higher SARS-CoV-2-specific antibodies in COVID-19 than non-COVID-19 respiratory samples.** **a** Heatmaps with unsupervised clustering of antibodies against SARS-CoV-2 and other human coronaviruses including SARS-CoV-1, 229E, NL63, OC43, and HKU1 in COVID-19 plasma and respiratory samples, as measured by multiplex bead array assay. **b** Median fluorescence intensity of IgG1-4 and antibodies with FcγR2aH, FcγR2aR, FcγR2b, FcγR3aV, FcγR3aF, or C1q binding abilities between COVID-19 and non-COVID-19 respiratory samples. Statistical significance was determined with Mann-Whitney test.

**Supplementary Fig. 3 Gating strategies for flow cytometry analyses. a** Respiratory myeloid antibody panel. **b** Whole blood lymphocyte antibody panel. **c** Whole blood innate T cell panel.

**Supplementary Fig. 4 Flow Self-Organizing Map (FlowSOM) analyses of respiratory samples. a**-**b** Cell surface marker expression of **a** respiratory myeloid antibody panel and **b** respiratory lymphocyte antibody panel. **c**-**d** Individual Fit-SNE plots of the **c** myeloid antibody panel and **d** lymphocyte antibody panel. **e** Representative flow cytometry plot of #059 blood neutrophils.

**Supplementary Table 1 DRASTIC cohort demographics**

| Patient | Age  range | Sex | Days post disease onset | | | | Days in hospital | Location during hospitalization | Oxygen supply | Drug therapy |
| --- | --- | --- | --- | --- | --- | --- | --- | --- | --- | --- |
|  |  |  | V1 | V3 | V5 | V7 |  |  |  |  |
| 001 | 71-80 | M |  | 19 |  |  | 30 | Ward | NP/HM | D+R |
| 002 | 41-50 | M |  |  | 8 |  | 9 | ICU | NP | D+R |
| 003 | 41-50 | M | 7 |  |  |  | 18 | ICU | ETT | D+R |
| 004 | 61-70 | F | 7 |  |  | 37 | 31 | ICU | ETT | D |
| 005 | 71-80 | F | 3 |  |  |  | 6 | Ward | NP | D+R |
| 006 | 31-40 | F | 11 |  |  | 12 | 2 | Ward | N | N |
| 007 | 21-30 | F | 7 |  |  | 8 | 2 | Ward | N | N |
| 008 | 21-30 | F | 11 |  |  | 12 | 2 | Ward | N | N |
| 009 | 51-60 | F | 7 |  |  | 8 | 2 | Ward | N | N |
| 010 | 51-60 | M | 11 |  |  |  | 6 | Ward | N | D |
| 011 | 41-50 | F | 7 |  |  |  | 27 | ICU | NP | D+R |
| 012 | 61-70 | M | 9 |  |  |  | 13 | ICU | NP | D+R |
| 013 | 61-70 | M | 10 |  |  |  | 12 | ICU | ETT | D+R |
| 014 | 31-40 | M | 10 |  |  |  | 6 | Ward | N | D |
| 015 | 21-30 | M | 5 |  |  |  | 2 | Ward | N | N |
| 016 | 61-70 | M | 12 |  |  |  | 9 | Ward | N | N |
| 017 | 71-80 | M | 13 |  |  |  | 4 | Ward | N | N |
| 018 | 71-80 | F | 7 |  |  |  | 4 | Ward | N | N |
| 019 | 21-30 | M | 5 |  |  | 16 | 11 | Ward | N | D+R |
| 020 | 71-80 | M | 5 |  |  |  | 9 | Ward | NP | D+R |
| 021 | 61-70 | F | 2 |  |  |  | 19 | Ward | NP/HM | D |
| 022 | 51-60 | M | 7 |  |  | 10 | 4 | Ward | N | N |
| 023 | 51-60 | M | 8 |  |  | 15 | 9 | ICU | NP/HM | D+R |
| 024 | 61-70 | F | 12 |  |  |  | 4 | Ward | N | N |
| 025 | 51-60 | F | 5 |  |  | 26 | 21 | ICU | HFNP | D |
| 026 | 51-60 | F | 16 |  |  | 36 | 29 | ICU | ETT | D |
| 027 | 51-60 | F | 6 |  |  |  | 3 | Ward | N | N |
| 028 | 31-40 | F | 12 |  |  | 17 | 6 | Ward | N | N |
| 029 | 61-70 | M | 7 |  |  | 28 | 23 | Ward | HFNP | D |
| 030 | 71-80 | F | 12 |  |  | 15/18^#^ | 47 | ICU | N | N |
| 031 | 81-90 | M | 2 |  |  | 10 | 24 | Ward | N | N |
| 032 | 31-40 | M | 11 |  |  |  | 1 | Ward | N | N |
| 033 | 21-30 | F | 9 |  |  |  | 1 | Ward | N | N |
| 034 | 41-50 | f | 8 |  |  | 11 | 4 | Ward | NP | D+R |
| 035 | 21-30 | F | 13 |  |  |  | 2 | Ward | N | D |
| 036 | 61-70 | M | 9 |  |  |  | 2 | Ward | N | N |
| 037 | 51-60 | F | 9 |  |  |  | 7 | Ward | N | D |
| 038 | 61-70 | F | 9 |  |  | 12 | 4 | Ward | NP | D |
| 039 | 81-90 | M | 9 |  |  | 14 | 5 | Ward | NP | D+R |
| 040 | 71-80 | M | 4 |  |  | 27 | 25 | Ward | NP | D |
| 041 | 31-40 | M | 6 |  |  |  | 1 | Ward | N | N |
| 042 | 21-30 | M | 7 |  |  | 12 | 4 | Ward | NP | D+R |
| 043 | 61-70 | M |  | 19 |  | 52 | 44 | ICU | ETT | D |
| 044 | 71-80 | F | 3 |  |  | 10/16^#^ | 17 | ICU | NP | D+R |
| 045 | 51-60 | M | 7 |  |  |  | 4 | ICU | NP | D+R |
| 046 | 81-90 | M | 15 |  |  | 24 | 11 | Ward | N | N |
| 047 | 71-80 | M | 8 |  |  |  | 6 | Ward | NP | D |
| 048 | 51-60 | M | 10 |  |  | 17 | 7 | ICU | NP | D |
| 049 | 21-30 | F | 7 |  |  |  | 6 | ICU | ETT | D+R |
| 050 | 51-60 | F | 7 |  |  |  | 12 | Ward | NP | D+R |
| 051 | 71-80 | F | 14 |  |  | 17 | 5 | Ward | NP | N |
| 052 | 51-60 | F | 11 |  |  | 16 | 6 | Ward | N | N |
| 053 | 81-90 | M | 6 |  |  | 15 | 13 | Ward | N | D |
| 054* | 41-50 | F | 3^ |  |  | 53 | 51 | ICU | ETT | N |
| 055* | 71-80 | F | 0^ |  |  | 19 | 19 | ICU | ETT | N |
| 056 | 51-60 | M | 35 |  |  | 41 | 8 | ICU | NP | D |
| 057 | 31-40 | F | 10 |  |  | 11 | 2 | Ward | N | N |
| 058 | 51-60 | M | 12 |  |  | 20 | 10 | Ward | NP | D |
| 059* | 81-90 | M | 7 |  |  |  | 22 | ICU | ETT | N |
| 060 | 41-50 | F | 13 |  |  | 16 | 4 | Ward | N | N |
| 061 | 71-80 | M | 0 |  |  |  | 7 | Ward | NP | D |
| 062* | 61-70 | M | 0^ |  |  |  | 13 | ICU | ETT | N |
| 063* | 81-90 | M | 3 |  |  |  | 10 | ICU | ETT | N |
| 064 | 41-50 | M |  |  |  |  | 7 | ICU | HFNP | D+R |
| 065 | 41-50 | F | 5 |  |  |  | 1 | Ward | N | N |
| 066* | 61-70 | F | 8 |  |  |  | 3 | Ward | NP | N |

*Non-COVID-19 patients with variable disease: 054, decompensated alcoholic hepatitis with encephalopathy; 055, hemangioblastoma intracranial haemorrhage; 059, Klebsiella pneumonia; 062, intracranial haemorrhage; 063, atypical pneumonia; 066, infective exacerbation of chronic obstructive pulmonary disease. ^Date of intubation was used because of no respiratory disease onset. ^#^Patients with delays after anticipated discharge date, data from the later sample were used for analyses if available. V1, hospital admission; V3, ARDS/CRS diagnosis; V5, 24-48 hours post drug therapy; V7, hospital discharge. Abbreviations: N, none; NP, nasal prong; NP/HM, nasal prong/ Hudson mask; HFNP, high flow nasal prong; D, dexamethasone; D+R, dexamethasone and remdesivir.

**Supplementary Table 2 Clinical summary of DRASTIC cohort**

| **COVID-19 positive patients** | **Total** (n=60) | **Ward** (n=43) | **ICU** (n=17) | **p value**^a^ | **COVID-19 negative patients** (n=6) | **p value**^b^ |
| --- | --- | --- | --- | --- | --- | --- |
| **Age** (years), median (range) | 58 (22-90) | 58 (22-90) | 58 (25-78) | 0.9256^c^ | 71 (41-85) | 0.0710^c^ |
| **Female**, n (%) | 28 (46.7%) | 21 (48.8%) | 7 (41.2%) | 0.7749^d^ | 3 (50%) | >0.9999^d^ |
| **Ethnicity**, n (%) |  |  |  | >0.9999^d, e^ |  | 0.6875^d, e^ |
| African | 3 (5%) | 1 (2.3%) | 2 (11.8%) |  | 0 |  |
| Arabic | 3 (5%) | 3 (7%) | 0 |  | 0 |  |
| Asian | 5 (8.3%) | 4 (9.3%) | 1 (5.9%) |  | 2 (33.3%) |  |
| Aboriginal and Torres Strait Islander | 1 (1.7%) | 1 (2.3%) | 0 |  | 0 |  |
| European | 33 (55%) | 24 (55.8%) | 9 (52.9%) |  | 4 (66.7%) |  |
| Indo-Asian | 2 (3.3%) | 2 (4.7%) | 0 |  | 0 |  |
| Middle Eastern | 7 (11.7%) | 5 (11.6%) | 2 (11.8%) |  | 0 |  |
| Pacific Islander | 1 (1.7%) | 0 | 1 (5.9%) |  | 0 |  |
| South Asian | 4 (6.7%) | 2 (4.7%) | 2 (11.8%) |  | 0 |  |
| Turkish | 1 (1.7%) | 1 (2.3%) | 0 |  | 0 |  |
| **Weight** (kg), median (range) | 77 (44.4-128) | 75.5 (44.6-110) | 88.7 (44.4-128) | 0.0102^c^ | 72 (45-91) | 0.3435^c^ |
| **Height** (cm), median (range) | 167 (152-193) | 165 (152-193) | 170 (157-185) | 0.1202^c^ | 164 (152-175) | 0.4404^c^ |
| **BMI** (kg/m^2^), median (range) | 26.7 (17.4-41.4) | 26.4 (17.4-41.4) | 30.4 (18-40.3) | 0.0653^c^ | 29 (18-32) | 0.6384^c^ |
| **Days from symptom onset to hospitalization**, median (range) | 7 (-1-48) | 7 (-1-14) | 7 (2-48) | 0.8994^c^ | 2 (0-7) | 0.0064^c^ |
| **Days in hospital**, median (range) | 6 (1-47) | 5 (1-30) | 13 (4-47) | <0.0001^c^ | 16 (3-51) | 0.0753^c^ |
| **Ward/ICU**, n (%) |  |  |  | N/A |  | 0.0136^d^ |
| Ward | 43 (71.7%) | N/A | N/A |  | 1 (16.7%) |  |
| ICU | 17 (28.3%) | N/A | N/A |  | 5 (83.3%) |  |
| **NIH score**, n (%) |  |  |  | 0.0001^d, f^ |  | N/A |
| 2 | 6 (10%) | 6 (14%) | 0 |  | N/A |  |
| 3 | 21 (35%) | 20 (46.5%) | 1 (5.9%) |  | N/A |  |
| 4 | 27 (45%) | 16 (5.3%) | 11 (64.7%) |  | N/A |  |
| 5 | 6 (10%) | 1 (2.3%) | 5 (29.4%) |  | N/A |  |
| **Oxygen support**, n (%) |  |  |  | <0.0001^d, g^ |  | 0.0308^d, g^ |
| **None** | 29 (48.3%) | 28 (65.1%) | 1 (5.9%) |  | 0 |  |
| **Non-Invasive** | 25 (41.7%) | 15 (34.9%) | 10 (58.8%) |  | 1 (16.7%) |  |
| Nasal prong | 19 (31.7%) | 12 (27.9%) | 7 (41.2%) |  | 1 (16.7%) |  |
| Nasal prong / Hudson mask | 3 (5%) | 2 (4.7%) | 1 (5.9%) |  | 0 |  |
| High flow nasal prong | 3 (5%) | 1 (2.3%) | 2 (11.8%) |  | 0 |  |
| **Invasive** | 6 (10%) | 0 | 6 (35.3%) |  | 5 (83.3%) |  |
| Endotracheal tube | 6 (10%) | 0 | 6 (35.3%) |  | 5 (83.3%) |  |
| **Clinical presentation**  (ILI/pneumonia/chest x-ray consolidation), n (%) | | |  | 0.3142^d^ |  | 0.1478^d^ |
| None | 13 (21.7%) | 11 (25.6%) | 2 (11.8%) |  | 3 (50%) |  |
| Yes | 47 (78.3%) | 32 (74.4%) | 15 (88.2%) |  | 3 (50%) |  |
| **Drugs**, n (%) |  |  |  | 0.0009^d, g^ |  | 0.0030^d, g^ |
| None | 24 (40%) | 23 (53.5%) | 1 (5.9%) |  | 6 (100%) |  |
| Dexamethasone (5-day course) | 18 (30%) | 12 (27.9%) | 6 (35.3%) |  | 0 |  |
| Dexamethasone (5-day course)  + Remdesivir (5-day course) | 18 (30%) | 8 (18.6%) | 10 (58.8%) |  | 0 |  |
| **Immunosuppressants**, n (%) |  |  |  | >0.9999^d, g^ |  | >0.9999^d, g^ |
| None | 52 (86.7) | 37 (86%) | 15 (88.2%) |  | 6 (100%) |  |
| Ciclosporin +  Mycophenolate mofetil | 1 (1.7%) | 1 (2.3%) | 0 |  | 0 |  |
| Vinblastine + Prednisolone +  Pembrolizumab | 1 (1.7%) | 1 (2.3%) | 0 |  | 0 |  |
| Methotrexate + Prednisolone | 1 (1.7%) | 0 | 1 (5.9%) |  | 0 |  |
| Prednisolone | 3 (5%) | 2 (4.7%) | 1 (5.9%) |  | 0 |  |
| Dexamethasone | 1 (1.7%) | 1 (2.3%) | 0 |  | 0 |  |
| Tacrolimus | 1 (1.7%) | 1 (2.3%) | 0 |  | 0 |  |
| Tacrolimus +  Mycophenolate mofetil | 0 | 0 | 0 |  | 0 |  |
| **Smoker**, n (%) |  |  |  | 0.6023^d, h^ |  | 0.0347^d, h^ |
| Non-smoker | 43 (71.7%) | 31 (72.1%) | 12 (70.6%) |  | 0 |  |
| Ex-smoker | 8 (13.3%) | 7 (16.3%) | 1 (5.9%) |  | 1 (16.7%) |  |
| Smoker | 5 (8.3%) | 3 (7%) | 2 (11.8%) |  | 2 (33.3%) |  |
| Unknown | 4 (6.7%) | 2 (4.7%) | 2 (11.8%) |  | 3 (50%) |  |

^a^Comparison between COVID-19 positive ward and ICU patients.

^b^Comparison between COVID-19 positive and COVID-19 negative patients.

^c^Significance was determined using the Mann-Whitney test.

^d^Significance was determined using the Fisher’s exact test.

^e^Comparison between European and other ethnicities (not including unknown).

^f^Comparison between NIH score 2, 3 and NIH score 4, 5.

^g^Comparison between patients with or without oxygen support, drug treatments, or immunosuppressants.

^h^Comparison between Non-smoker and Ex-smoker, and Smoker (not including unknown).

Abbreviations: ICU, intensive care unit; N/A, not applicable; NIH, National Institutes of Health; ILI, influenza-like-illness.

**Supplementary Table 3 Grading criteria of the National Institutes of Health (NIH) score***

| **NIH score** | **Criteria** |
| --- | --- |
| Asymptomatic/ Presymptomatic (1) | Individuals who:   - test positive for SARS-CoV-2 by virologic testing, and - have no symptoms |
| Mild (2) | Individuals who:   - have COVID-19 symptoms such as fever, cough, sore throat, malaise, headache, muscle pain, and - without shortness of breath, dyspnoea, or abnormal chest imaging |
| Moderate (3) | Individuals who:   - have evidence of lower respiratory disease by clinical assessment or imaging, and - have a saturation of oxygen (SpO_2_) ≥94% on room air at sea level |
| Severe (4) | Individuals who:   - have respiratory frequency >30 breaths per minute, or - SpO_2_ <94% on room air at sea level, or - ratio of arterial partial pressure of oxygen to fraction of inspired oxygen (PaO_2_/FiO_2_) <300 mmHg, or - lung infiltrates >50% |
| Critical (5) | Individuals who:   - have respiratory failure, septic shock, and/or multiple organ dysfunction |

*https://www.covid19treatmentguidelines.nih.gov/overview/clinical-spectrum/

**Supplementary Table 4 Validation test of the surrogate Virus Neutralization Test**

| **Sample** | **COVID-19 status** | **Dilution**^a^ | **% Inhibition**^b^ |
| --- | --- | --- | --- |
| COVID-19 positive serum | Positive | Neat | 95.6 |
|  |  | 1:2 | 86.4 |
|  |  | 1:10 | 44.1 |
| DRASTIC 055 ETA | Negative | Neat | 10.5 |
|  |  | 1:2 | 87.3 |
|  |  | 1:10 | 43.1 |
| DRASTIC 066 Sputum | Negative | Neat | 0 |
|  |  | 1:2 | 86.9 |
|  |  | 1:10 | 40.1 |

^a^COVID-19 negative ETA and sputum were tested in neat or mixed with the COVID-19 positive serum until the COVID-19 positive serum was in the dilution as stated in the column.

^b^Positive % inhibition was defined as ≥ 20%.

Abbreviation: ETA, endotracheal aspirate.

**Supplementary Table 5 Panel design of the multiplex bead array assay**

| **Pathogens** | **Proteins** | **Isotypes and FcɣR/C1q bindings** |
| --- | --- | --- |
| SARS-CoV-2 | RBD | IgG |
|  | S1 | IgG1 |
|  | S2 | IgG2 |
|  | Trimeric S | IgG3 |
|  | NP | IgG4 |
| SARS-CoV-1 | S1 | IgA1 |
|  | Trimeric S | IgA2 |
|  | NP | IgM |
| HCoV 229E | S1 | FcɣR2aH |
|  | S1+2 | FcɣR2aR |
|  | NP | FcɣR2b |
| HCoV NL63 | S1 | FcɣR3aV |
|  | S1+2 | FcɣR3aF |
|  | Trimeric S | C1q^a^ |
|  | NP |  |
| HCoV OC43 | S1 |  |
|  | S2 |  |
|  | S1+2 |  |
|  | NP |  |
| HCoV HKU1 | S1 |  |
|  | S1+2 |  |
|  | Trimeric S |  |
|  | NP |  |
| C. Tetani | Tetanus Toxin |  |
| Influenza A/Cali/07/2009 (H1N1) | Hemagglutinin |  |

^a^C1q binding was not tested for NP of SARS-CoV-2, SARS-CoV-1, HCoV 229E, HCoV NL63, HCoV OC43 and HCoV HKU1, and HCoV 229E S1+2. Abbreviations: FcɣR, fragment crystallizable region gamma receptor; C1q, complement component 1q; RBD, receptor binding domain; S, spike; NP, nucleoprotein; HCoV, human coronavirus.

**Supplementary Table 6 Flow cytometry antibody panels**

| **Colour** | **Fluorochrome** | **Respiratory myeloid panel** | **Respiratory lymphocyte panel** | **Whole blood lymphocyte panel** | **Whole blood innate T cell panel** |
| --- | --- | --- | --- | --- | --- |
| Violet | BV421 | CD66b | CXCR5 | CD71 |  |
|  | BV510 | CD64 |  | CD19 | Live/Dead |
|  | BV605 | HLA-DR | HLA-DR | HLA-DR | CD161 |
|  | BV650 | CD4 | CD4 | CD4 |  |
|  | BV711 | CD32 | CD27 | CD27 | TRAV1-2 |
|  | BV786 | CD11b | CD38 | CD38 |  |
| Red | APC | CD62L | CD56 | CD56 |  |
|  | AF700 | CD16 | CD16 | CD16 | CD27 |
|  | APC-H7 | Live/Dead | Live/Dead | CD14 | CD19 |
| Blue | FITC |  | CD45RA | CD45RA | TCRgd |
|  | PerCP-Cy5.5 | CD45 | CD45 | CD8 |  |
| Yellow-Green | PE | CD38 | TCRgd | TCRgd | MR1-5-OP-RU tetramer |
|  | ECD | CD19 | CD19 | CD3 |  |
|  | PE-Cy7 | CD1c | PD-1 |  |  |
| UV | BUV395 | CD3 | CD3 |  | CD3 |
|  | BUV496 |  |  |  | CD4 |
|  | BUV737 | CD14 | CD14 |  |  |
|  | BUV805 | CD8 | CD8 |  | CD8 |
